# Molnupiravir clinical trial simulation suggests that polymerase chain reaction underestimates antiviral potency against SARS-CoV-2

**DOI:** 10.1101/2024.11.21.24317726

**Authors:** Shadisadat Esmaeili, Katherine Owens, Joseph F. Standing, David M. Lowe, Shengyuan Zhang, James A. Watson, William H. K. Schilling, Jessica Wagoner, Stephen J. Polyak, Joshua T. Schiffer

## Abstract

Molnupiravir is an antiviral medicine that induces lethal copying errors during SARS-CoV-2 RNA replication. Molnupiravir reduced hospitalization in one pivotal trial by 50% and had variable effects on reducing viral RNA levels in three separate trials. We used mathematical models to simulate these trials and closely recapitulated their virologic outcomes. Model simulations suggest lower antiviral potency against pre-omicron SARS-CoV-2 variants than against omicron. We estimate that in vitro assays underestimate in vivo potency 7-8 fold against omicron variants. Our model suggests that because polymerase chain reaction detects molnupiravir mutated variants, the true reduction in non-mutated viral RNA is underestimated by ∼0.5 log_10_ in the two trials conducted while omicron variants dominated. Viral area under the curve estimates differ significantly between non-mutated and mutated viral RNA. Our results reinforce past work suggesting that in vitro assays are unreliable for estimating in vivo antiviral drug potency and suggest that virologic endpoints for respiratory virus clinical trials should be catered to the drug mechanism of action.

## Introduction

Molnupiravir is an antiviral prodrug that induces errors in the SARS-CoV-2 genome, which typically renders the virus unable to replicate further.^1^ In the randomized double-blinded MOVe-OUT trial, which enrolled unvaccinated individuals when delta, mu, and gamma variants of concern (VOC) were circulating, molnupiravir reduced hospitalization by 50% and viral load after treatment (day 5) by 0.3 log relative to placebo.^2^ In the platform adaptive PANORAMIC trial, which enrolled vaccinated individuals when omicron VOCs were circulating, hospitalization rates were only 1% in both arms, but molnupiravir lowered viral load after treatment by 0.94 log_10_ relative to usual care.^3^ In the adaptive platform trial PLATCOV, which enrolled low-risk individuals when omicron VOCs were circulating, molnupiravir lowered viral load after treatment by 1.09 log relative to usual care.^4^ Taken together, these trials demonstrate that molnupiravir has both clinical and virologic efficacy which varied across trials and viral variants.

Overall, use of molnupiravir has been lower than that of nirmatrelvir / ritonavir based on lower reduction in hospitalization in MOVe-OUT relative to the EPIC-HR trial for nirmatrelvir / ritonavir.^5^ A concern has also been raised that molnupiravir’s mechanism of action could generate novel mutants that persist after cessation of treatment,^6^ and then spread in the population.^7^ Nevertheless, the PANORAMIC and PLATCOV trial results suggest high potency, and molnupiravir is still considered in individuals in whom nirmatrelvir / ritonavir is contraindicated and in combination with other drugs in immunocompromised hosts.^8, 9^ There is currently no explanation for the disparate antiviral effects in MOVe-OUT versus PANORAMIC and PLATCOV. Moreover, the fact that polymerase chain reaction (PCR) detects drug-altered viral RNA molecules^6^ has not been considered in the analysis of trial outcomes. A study in ferrets highlights the potential importance of this effect: while molnupiravir and nirmatrelvir / ritonavir dramatically lowered levels of infectious SARA-CoV-2 titers, only nirmatrelvir / ritonavir lowered total viral RNA levels.^10^

We previously used clinical trial simulation to reproduce results from nirmatrelvir / ritonavir trials for SARS-CoV-2.^11, 12^ We first validated a viral immune dynamic model (VID) against a very large prospective cohort of infections that included multiple VOCs.^12^ We used sets of model parameters that reflect diverse virologic output to create simulated cohorts for the control arms of trials. We then integrated pharmacokinetic (PK) and pharmacodynamic (PD) models for nirmatrelvir / ritonavir with the VID models to simulate treatment arms.^11^ This approach recapitulated mean viral load reduction in the EPIC-HR and PLATCOV trials, as well as individual viral load trajectories in PLATCOV. The validated model was then used to explain the high frequency of virologic and concurrent symptomatic rebound with use in the community,^13^ despite very low levels of virologic and symptomatic rebound in the EPIC-HR trial.^14, 15^ Model output suggests that extending therapy from 5 to 10 days would nearly eliminate rebound,^11^ a result confirmed with modeling of separate data.^16^

Here we expand this approach to develop a new joint VID-PK-PD model to account for the unique mechanism of action of molnupiravir. We fit the model to results from the MOVe-OUT, PLATCOV and PANORAMIC trials. Our results suggest that quantitative viral PCR likely underestimates the reduction in non-mutated viral RNA and therefore the true potency of molnupiravir during omicron infections.

## Results

### Viral immune dynamic, pharmacokinetic and pharmacodynamic clinical trial simulation models

We previously described our viral immune dynamic (VID) model that was fit to diverse serial viral loads from 1510 SARS-CoV-2 infected individuals in the National Basketball Association cohort.^12, 17^ The model assumes a finite number of susceptible cells and an eclipse phase delays viral production by infected cells. In keeping with an early innate immune response, susceptible cells become refractory to infection in the presence of infected cells but also revert to a susceptible state at a constant rate. Infected cells are cleared by cytolysis and delayed acquired immunity, which is activated in a time-dependent fashion **(Fig 1a)**. We used a mixed-effect population approach implemented in Monolix to estimate model parameters.^18^

**Fig 1.**
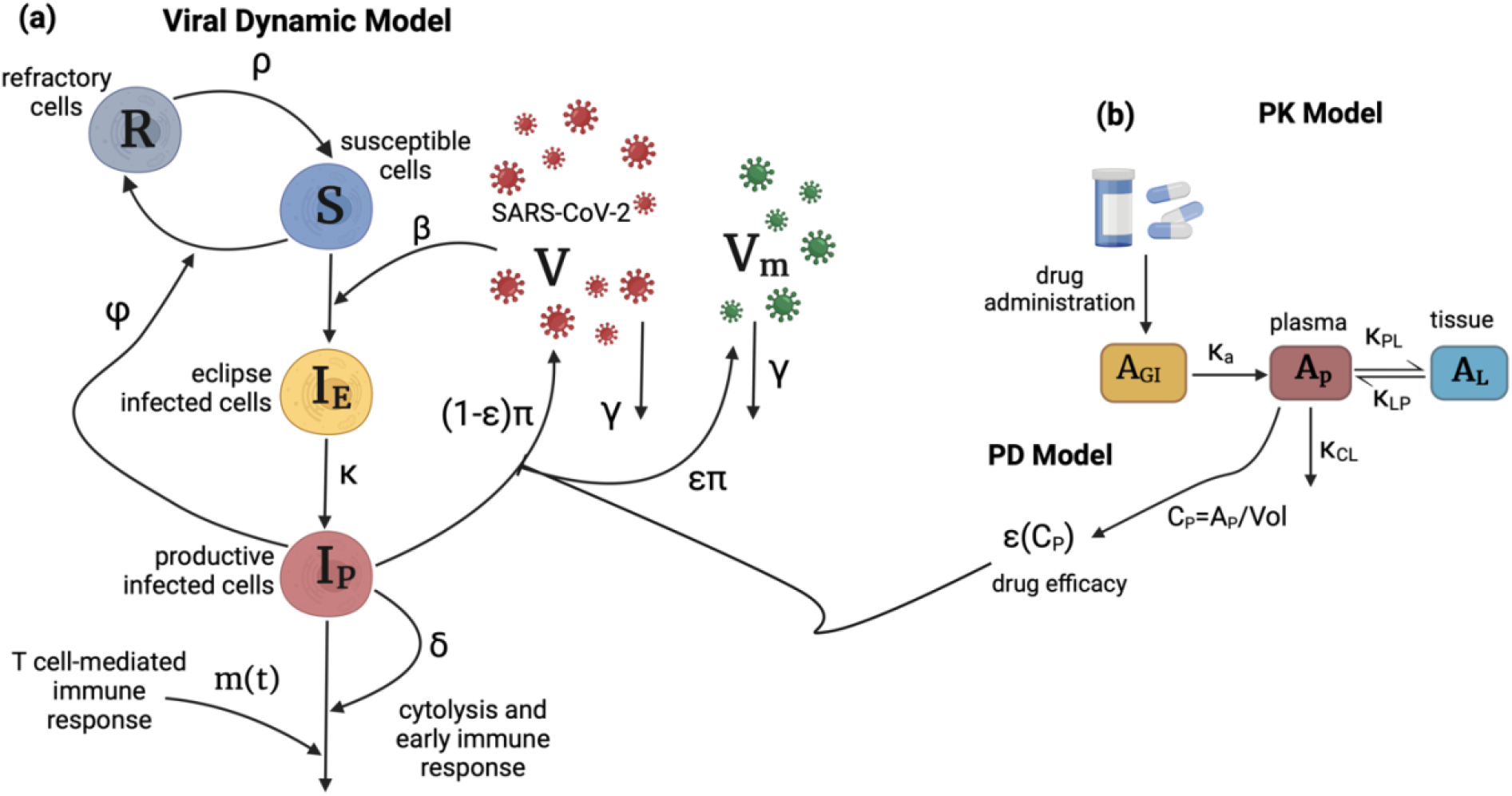
Schematic of the viral dynamic model and molnupiravir PK-PD model. **(a)** In the viral dynamic model, S represents the susceptible cells, *I*_*E*_ is the eclipse infected cells, *I*_*P*_ is the productively infected cells, V is the non-mutated viruses, and V_m_ is the mutated viruses as a result of treatment. The productively infected cells are cleared by early and late T cell-mediated immune response at rates *δ* and *m*(*t*). *β* is the infectivity rate, *ϕ* is the rate of conversion of susceptible cells to refractory cells, and *ρ* is the rate of reversion of the refractory cells to susceptible cells. productively infected cells produce viruses at the rate *π*, and free viruses are cleared at the rate *γ*. **(b)** Two-compartmental PK model with oral administration of the drug which models the amounts of the drug in gut tissue (A_GI_), plasma (A_P_), and the respiratory tract (A_L_). *κ*_*a*_ is the rate of absorption of the drug from gut to plasma, *κ*_*PL*_ and *κ*_*LP*_ are the rates of transfer of the drug from plasma to the respiratory tract and back, and *k*_*CL*_is the rate at which the drug clears from the body. Vol is the estimated plasma volume and *C*_*P*_ is the concentration of the drug in plasma. *ϵ*(*C*_*P*_) is the efficacy of the drug in converting produced viruses into mutated viruses.

To reproduce levels of molnupiravir, we used a two-compartment pharmacokinetic (PK) model (**Fig. 1b**). Using Monolix and the mixed-effect population approach, we estimated parameter values by fitting the model to the average plasma concentration of healthy subjects.^19^ The model closely recapitulated observed drug levels following multiple doses of 50, 100, 200, 300, 400, 600, and 800 mg given twice daily for five days (**Fig S1, Table S1**). The estimated value for the transition rate from plasma to peripheral compartment (*k*_*PL*_) was dose-dependent in the form of *k*_*PL*_ = *k*_*PL*,1_*Dose*^*α*^. All other PK parameters were dose independent. For the pharmacodynamic (PD) model, we assumed drug efficacy follows a Hill equation with respect to concentration. We parameterized the model using in vitro efficacy data collected at different concentrations (details in **Materials and Methods, Fig S2, Table S2**).^20^

We combined the VID and PKPD models by using treatment efficacy to convert non-mutated virus to mutated virus, both of which are assumed to be detected with polymerase chain reaction (PCR) assays, given the low probability of drug-induced mutations in the PCR primer region. This assumption is based on the observed drug-induced mutation rate of approximately 1 mutation per 2000 base pairs.^21^ Given the average length of most PCR primers of ∼25 base pairs, the chance of the primer remaining unmutated after treatment is (1999/2000)^25^, or 98.76%. A limitation of viral load data from the included clinical trials is that it lacks early pre-symptomatic endpoints to estimate the viral expansion slope. To further train the model, we included omicron-infected participants from the NBA cohort (n=1023) to inform rates of viral upslope in the trials.^12^ We first fit the combined model to individual viral load data from 149 low-risk, symptomatic vaccinated participants infected with omicron VOCs in the PLATCOV trial (65 treated and 84 controls) and from 80 high-risk, symptomatic vaccinated participants infected with omicron VOCs in the PANORAMIC trial (38 treated and 42 controls) (**Fig 2a, 2b, Fig S3, S4, S5, S6**). We next fit the combined model to trial endpoint data (mean viral load drop from baseline) reported in three randomized, controlled trials: PLATCOV (**Fig 3a, 4a-d**),^4^ PANORAMIC (**Fig 3b, 4e-h**),^3^ and the MOVe-OUT trial with 1093 high-risk unvaccinated symptomatic individuals infected with pre-omicron VOCs (549 treated + 544 placebo, **Fig 3c, 4i-k**).^2^ All model fitting was performed using Monolix with non-linear mixed effects approaches described in the **Materials and Methods**.

**Fig 2.**
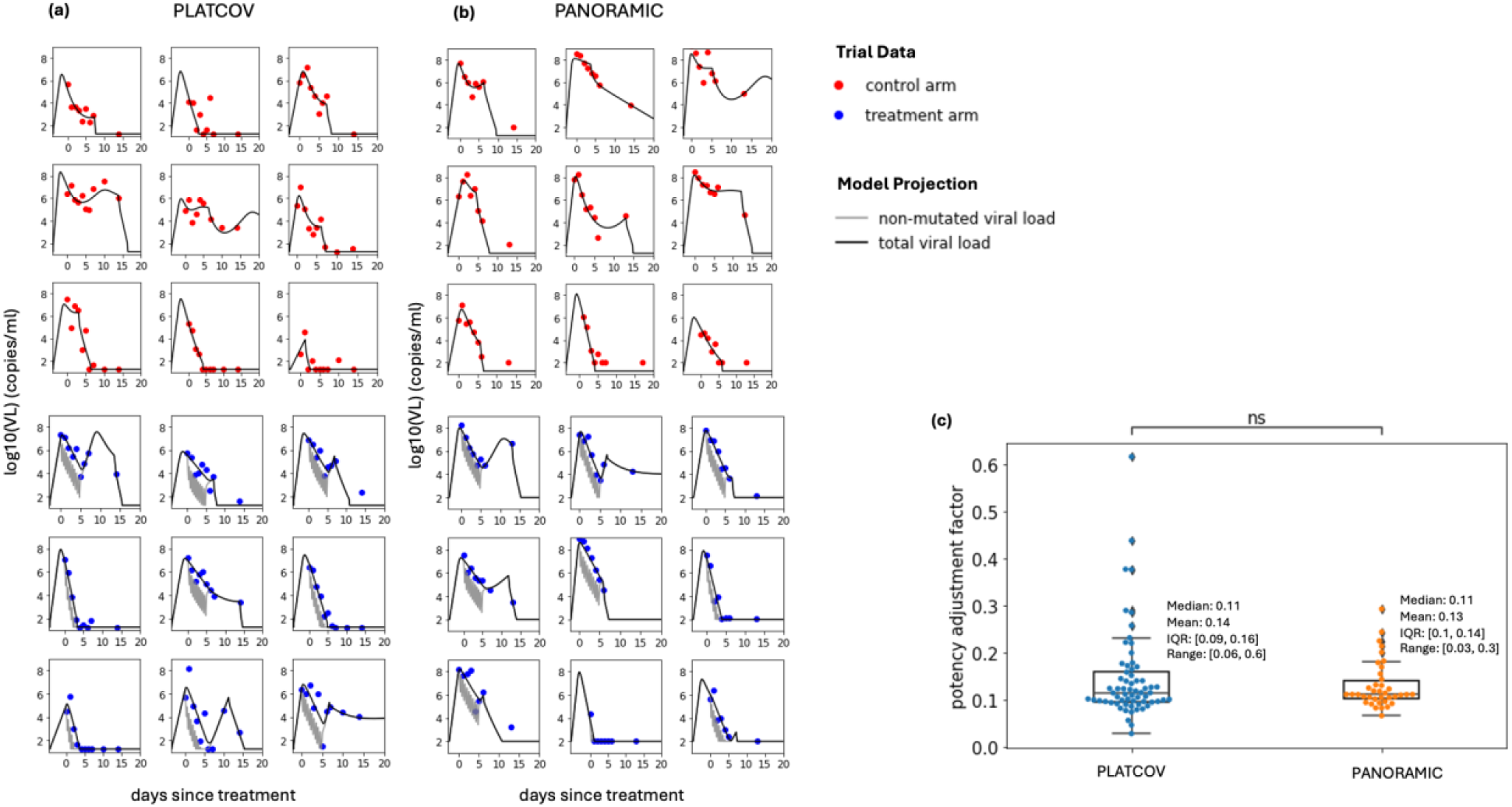
Mathematical model fits of SARS-CoV-2 viral load over time to a subset of study participants in PLATCOV and PANORAMIC receiving no treatment (control) or molnupiravir. **(a)** Model fits to 9 control and 9 treatment participants in PLATCOV. **(b)** Model fits to 9 control and 9 treatment participants in PANORAMIC. **(c)** Individual estimates for potency adjustment factor (ratio of *in vivo*: *in vitro* EC50) in the two trials (center line, median; box limits, upper and lower quartiles; whiskers, 1.5x interquartile range). The statistical comparison was performed using Mann-Whitney U-test.

**Figure 3.**
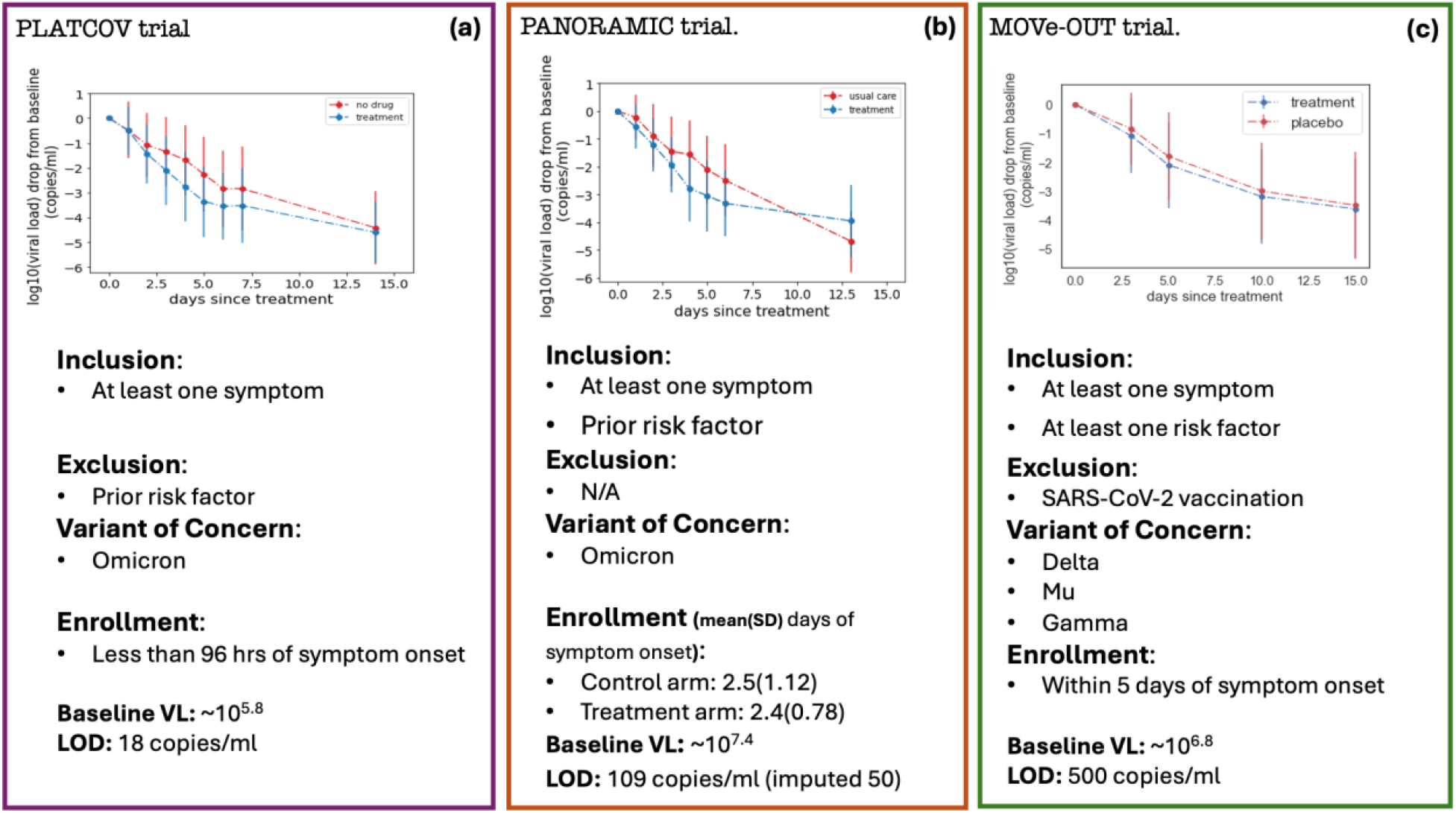
Mean viral load reduction in three trials which are targets for model fitting: **(a)** PLATCOV, including individuals with no prior risk factor infected by omicron variant, and enrolled within 96 hours of symptom onset, **(b)** PANORAMIC, including individuals with prior risk factor infected by omicron variant, and enrolled on average 2.5 days since symptom onset, and **(c)** MOVe-OUT, including high-risk, unvaccinated individuals, infected by delta, mu, and gamma variants, and enrolled within five days of symptom onset.

### Model fitting to individual viral load trajectories in PLATCOV and PANORAMIC

For each participant, we defined the in vivo EC_50_ as the plasma drug concentration required to inhibit viral replication by 50% and the potency adjustment factor (paf) as the ratio between the in vivo and in vitro EC_50_.^22, 23^ To estimate the paf, we fit the combined VID-PKPD model to individual viral load data from both arms of the PLATCOV and PANORAMIC trials, as well as omicron infected individuals in the NBA cohort using the population mixed effect approach in Monolix. We achieved good model fit to individual viral load trajectories in the control and treatment arms of PLATCOV **(Fig 2a, Fig S3, S4)** and PANORAMIC **(Fig 2b, S5, S6**). The model projected higher levels of total detected SARS-CoV-2 RNA in most participants relative to non-mutated viral RNA **(Fig 2a,b)**. We estimated a range of individual paf values with similar mean and median values estimated for both trials **(Fig 2c**). These values suggest that in vivo potency of molnupiravir is on average 7-8 fold higher than estimates based on in vitro data. Each participant had an estimated paf less than one indicating that enhanced potency in vivo is necessary to accurately model the data.

### Model fit to trial virologic endpoint data from PLATCOV, PANORAMIC, and MOVe-OUT

As a second approach, we assessed whether a virtual cohort strategy where control participants are modeled using estimated parameter values from pre-existing cohorts can predict virologic trial endpoints. This approach is necessary for situations where individual viral load data are not available as with MOVe-OUT and demonstrates that the model can reproduce the primary virologic endpoint of each study. We simulated virtual cohorts using the combined VID-PKPD model and fit results to viral load decay from baseline in the three trials. For each trial arm, we randomly selected 400 individuals from the NBA cohort with the closest matching viral variant, symptom, and vaccine status and used their estimated individual viral dynamic parameters in simulations. To address variability in timing of baseline viral load measurement relative to infection, we randomly assigned all individuals an incubation period selected from a variant-specific gamma distribution found in the literature.^24, 25^ Treatment start day was randomly selected from a distribution based on observed enrollment windows in the three trials. Due to the lack of individual PK data, the same estimated population PK parameters were used for all simulated treated individuals (**Table S1)**. PD parameters were randomly selected from a log-normal distribution with estimated mean and standard error from in vitro assay results (**Table S2**).

Our model closely reproduced kinetics of viral decay in the PLATCOV control **(Fig 3a, 4a)** and treatment arms **(Fig 3a, 4b)** and estimated a paf=0.13 **(Fig 4c)** similar to our mean estimate using individual fits **(Fig 2c)**. The model also predicted individual-level variability in virologic responses observed in PLATCOV, including instances of increased viral load following therapy **(Fig 4d)**. We compared simulated and actual distributions of viral load change among trial participants in the control and treatment arms. On most post-treatment days, simulated and actual distributions were not statistically dissimilar. Wider distributions of observed versus simulated viral load change were noted on post-randomization days 1 and 2 for control and treatment **(Fig 4d)**, likely due to noise in viral load data from oral swabs: differences of 1–2 logs were often noted between replicates collected from PLATCOV participants at equivalent timepoints, particularly on days 1 and 2.^11^

**Figure 4.**
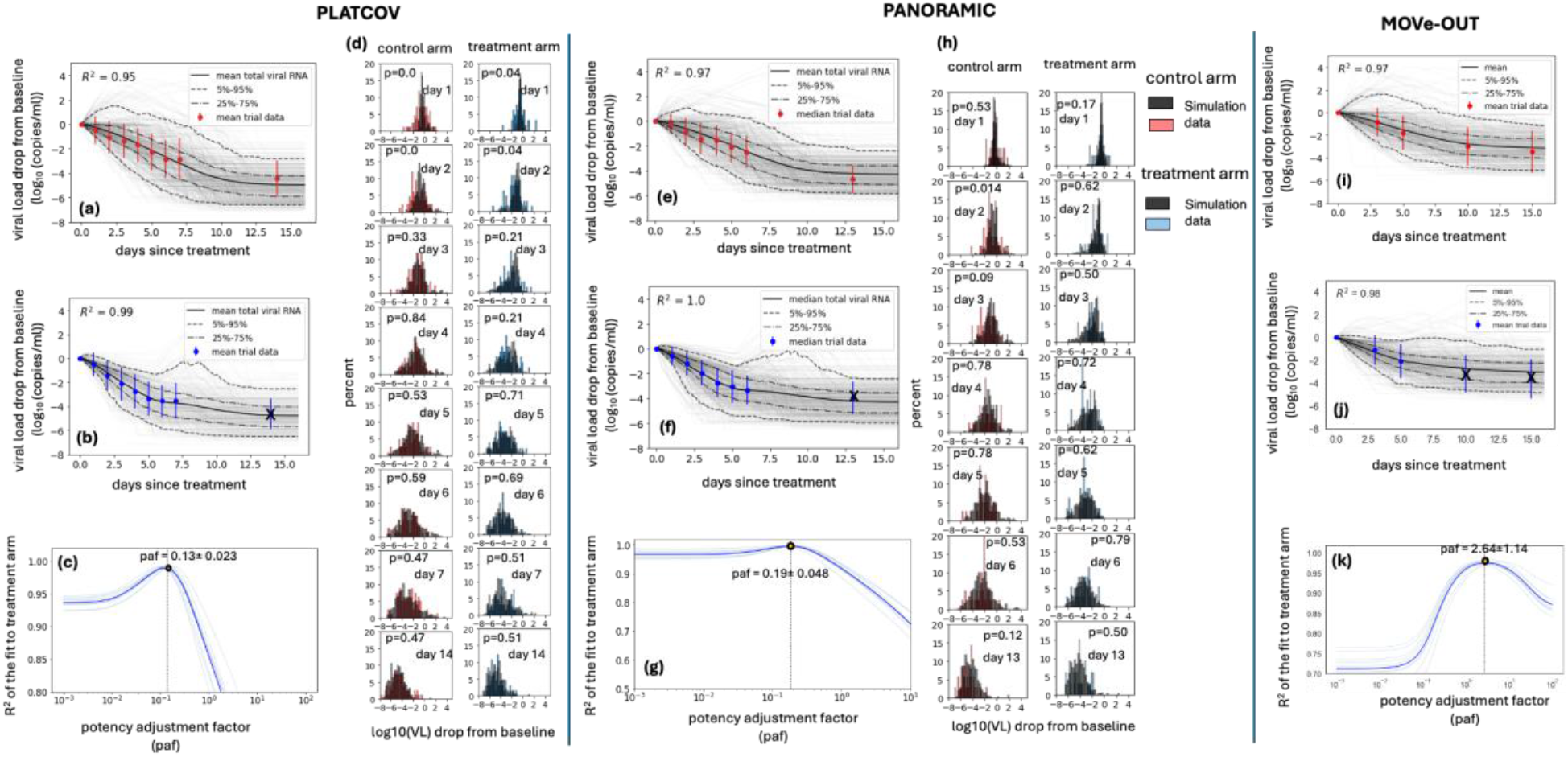
Model fit to virologic trial outcomes for (a-d) PLATCOV, (e-h) PANORAMIC, (i-k) MOVe-OUT. Results include **(a, e, i)** control groups, **(b, f, j)** treatment groups, **(d**,**h)** comparing individual variability of data vs simulation in control and treatment arms, and **(c, g, k)** estimate for potency adjustment factor. To only capture the effect of treatment and address potential identifiability issues, data from the first seven days after baseline were used to estimate the paf. Therefore, the crossed-out data points were not included in the calculation of the R^2^.

Similarly, our model closely reproduced kinetics of viral decay in PANORAMIC in control **(Fig 3b, 4e)** and treatment arms **(Fig 3b, 4f)** and estimated a paf=0.19 **(Fig 4g)** similar to our mean estimate using individual fits **(Fig 2c)**. The model also predicted individual-level variability in virologic responses observed in PANORAMIC, including instances of increased viral load following therapy **(Fig 4h)**. We compared simulated and actual distributions of viral load change among trial participants in the control and treatment arms. On all post-treatment days other than day 2 control, simulated and actual distributions were not statistically dissimilar **(Fig 4h)**. This likely indicates less noise in viral load data from nasopharyngeal swabs collected in PANORAMIC relative to oral swabs in PLATCOV.

Finally, the model reproduced kinetics of viral decay in MOVe-OUT in control **(Fig 4i)** and treatment arms **(Fig 4j)** but estimated a higher paf=2.64 **(Fig 4k)**. The higher paf maps to the far less substantial viral load reduction in MOVe-OUT relative to the other two trials which in turn might be explained by less potency against pre-omicron variants which has been observed experimentally.^26^

As a further validation step, we performed counterfactual simulations which assess viral kinetics of control study participants assuming treatment and treatment participants assuming placebo/usual care. Counterfactual control simulations slightly overestimated late viral loads for PLATCOV **(Fig S7a)** and PANORAMIC **(Fig S8a)**. This may be because therapy suppresses acquired immune responses, which is not captured in our model.^16^ Counterfactual treatment simulations fit the data well for PLATCOV **(Fig S7b)** and PANORAMIC **(Fig S8b)**. Simulations occasionally predicted viral rebound following treatment **(Fig S7c**,**d and S8c**,**d)**.

### Estimates of reduction in fully mutated viruses versus non-mutated SARS-CoV-2 RNA

We used our optimized model with solved paf to project the trajectory of non-mutated viral RNA during treatment relative to values measured with PCR, which detects viral RNA with drug-induced mutations.^6^ In PLATCOV **(Fig 5a,d,f)** and PANORAMIC **(Fig 5b,d,f)**, owing to higher drug potency, total SARS-CoV-2 viral RNA on treatment exceeded non-mutated viral RNA by ∼0.48 and 0.59 log respectively, suggesting that measured endpoints underestimate the drug’s true antiviral effect. However, these differences did not achieve statistical significance, perhaps because estimated total and non-mutated viral RNA levels converge at drug trough. In MOVe- OUT **(Fig 5c,d,f)**, there was no significant difference between total SARS-CoV-2 viral RNA on treatment and non-mutated viral RNA.

**Figure 5.**
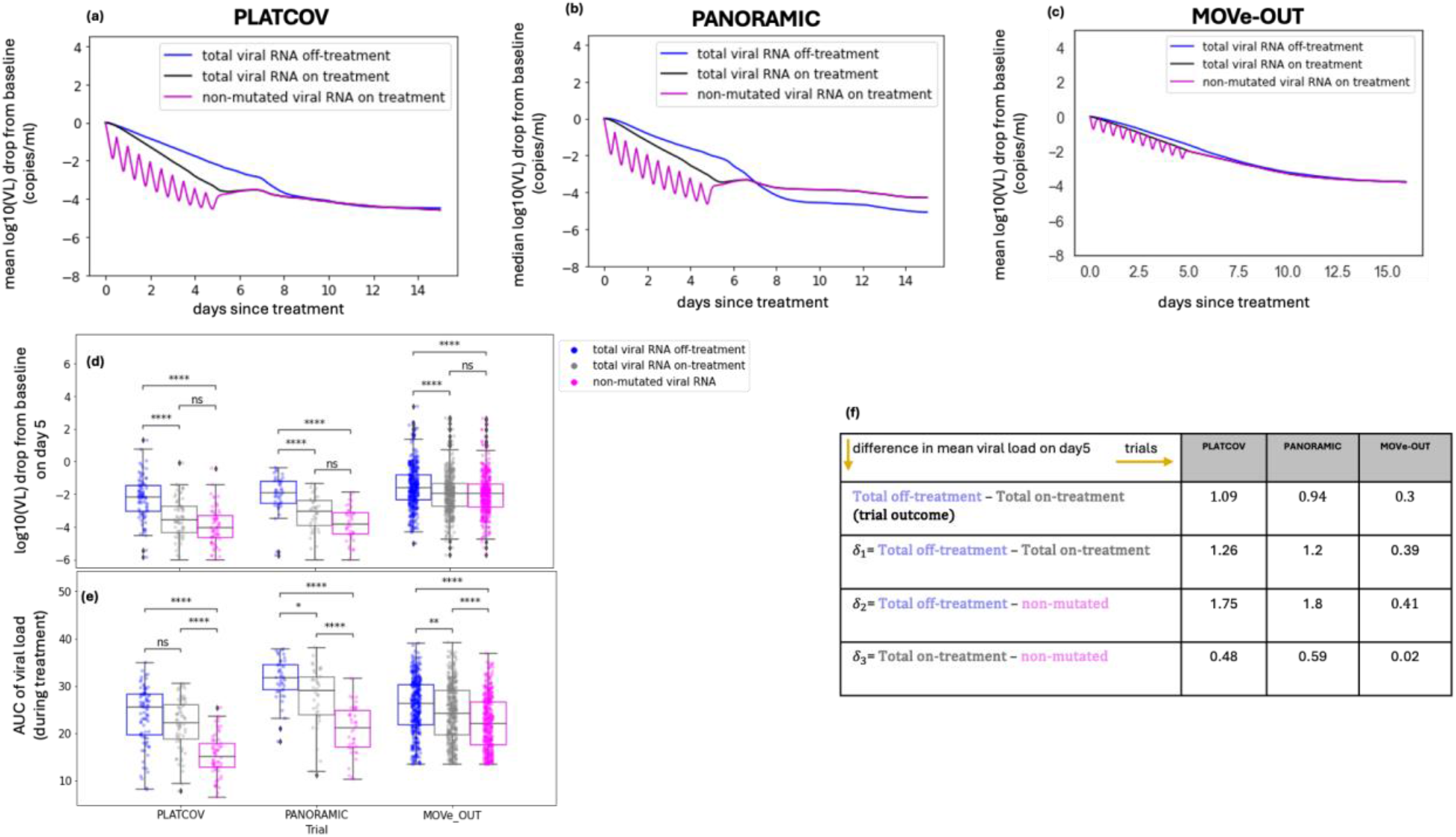
PCR underestimates the true reduction in non-mutated SARS-CoV-2 RNA in PLATCOV and PANORAMIC. Simulated mean viral loads including non-mutated viral RNA in (a) PLATCOV, (b) PANORAMIC and (c) MOVe-OUT. (d) Individual reduction at day 5 in the simulated control group (blue), simulated total viral RNA (grey) and simulated non-mutated viral RNA (pink) in the three trials showing no statistical difference between total and non-mutated viral RNA despite a lower median. (e) Individual viral area under the curve from the start of the treatment through day 5 in the simulated control group (blue), simulated total viral RNA (grey), and simulated non-mutated viral RNA (pink) in the three trials showing a statistical difference between total and non-mutated viral RNA. Boxplots include the interquartile range (IQR) with whiskers equaling 1.5 the IQR. (f) Table of mean viral load reductions in the trials and simulations including the predicted mean difference in total versus mutated viral RNA.

In all 3 trials, the models suggest that non-mutated viral loads during treatment may be lowest at drug peak, reflecting the short plasma half-life of molnupiravir. Therefore, the cumulative effect of drug is best estimated with viral area under the curve (AUC) which accounts for highly variable drug activity over time due to short drug half-life. By this estimate, the reduction in non-mutated viral RNA far exceeds that of total measured viral RNA **(Fig 5e)**. In the case of MOVe-OUT, this may explain why significant clinical benefit is associated with only a marginal reduction in observed decline in total viral RNA. This also suggests that there might be utility to measure viral load after drug peak and trough for agents with short half-life as viral loads may differ by a full order of magnitude according to drug level.

To mimic endpoints in PLATCOV, we estimated the slope of viral decay of the total viral RNA on and off-treatment as well as the non-mutated viral RNA by fitting a line to simulated data on days 0-5 after the start of the treatment. The median clearance half-life in our simulations of PLATCOV control and treatment arms reflected those observed in the trial **(Table S3)**. Our results showed small differences (non-significant) between the clearance rates for the total versus the non-mutated viral RNA in the treated groups in PLATCOV and PANORAMIC (**Fig S9**).

### Differences in trial participants and model parameters

We also compared features of each trial as they related to model predictions by assessing the viral dynamic range in each trial **(Fig S10a)**. Control participants in PLATCOV had lower mean viral loads throughout the course of infection relative to PANORAMIC and MOVe-OUT **(Fig S10b)**. Given PLATCOV and PANORAMIC enrolled participants with the omicron variant, we surmise that these differences relate to demographic differences in study participant demographics, slightly shorter estimated time to treatment in PANORAMIC versus PLATCOV estimated by our model (t_0_ in **Fig S11**), and/or characteristics of the PCR assay used in the studies. The trials also employed different limits of detection which impacted observed reductions in viral load **(Fig S10b)**. Model parameters were largely equivalent between studies and between treatment and control arms, reflecting the flexibility of the model **(Fig S11)**. The parameter governing transition of susceptible cells to a refractory state was higher in PLATCOV relative to PANORAMIC which likely was necessary to achieve lower viral loads overall and may reflect the younger study participants **(Fig S11)**.

### Antiviral potency, viral load assessment and trial design all impact observed antiviral reduction

We next combined PK and PD models to assess the average efficacy of the drug during days 0-5 in all three trials **(Fig 6a,b)** and noted an efficacy of 53% in MOVe-OUT **(Fig 6c)**. The efficacy of molnupiravir in PLATCOV (94%) and PANORAMIC (95%) was similar to that of nirmatrelvir in PLATCOV (94%) and EPIC-HR (82%) owing to a much lower paf for molnupiravir relative to nirmatrelvir / ritonavir **(Fig 6c)**.

**Figure 6.**
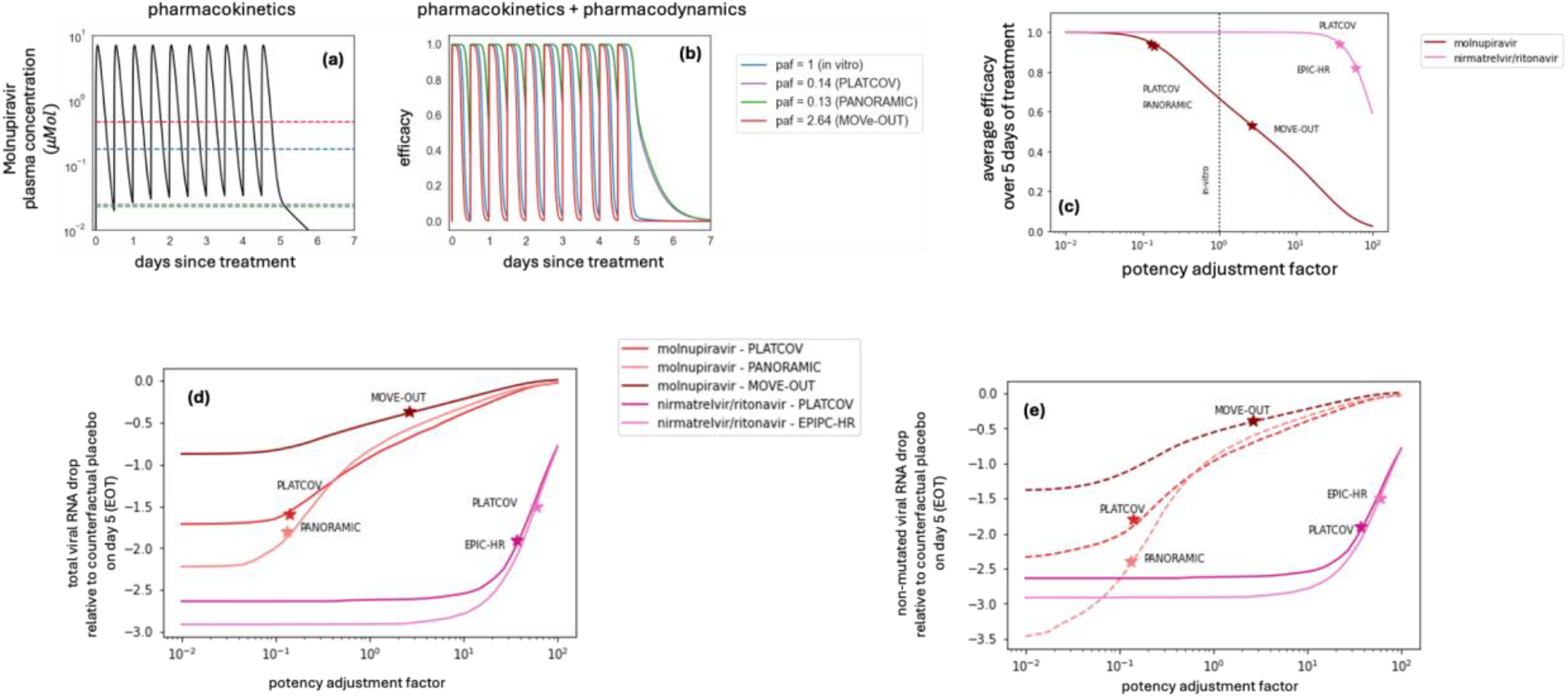
Relationship of drug pharmacokinetics and pharmacodynamics to in vivo potency and viral load reduction, comparing trial design. (a) Molnupiravir plasma concentration during five days of treatment with 800 mg molnupirvir given twice daily. The dashed lines mark the EC_50_ with different paf values which differ by trial. For paf = 0.13, drug levels are almost entirely above the EC_50_. (b) Dynamic shifts in molnupiravir efficacy for different paf values which differ by trial. Efficacy only drops minimally at trough levels when paf is low (i.e. 0.14 and 0.13 in PLATCOV and PANORAMIC) but drops significantly at trough levels in MOVe-OUT. (c) Drug potency of SARS-CoV-2 antivirals according to trial. The in-vivo efficacy of molnupiravir in PLATCOV and PANORAMIC trials is close to the in vivo efficacy of nirmatrelvir/ritonavir in the PLATCOV trial and higher than EPIC-HR. MOVe-OUT potency is significantly lower due to a higher paf and higher in vivo EC50 value. (d) Simulated mean drops in total viral RNA from baseline relative to counterfactual placebo/usual care arms on day 5 in the three molnupiravir trials and two nirmatrelvir/ritonavir trials. (e) Simulated mean drops in non-mutated viral RNA from baseline relative to counterfactual placebo on day 5 in the three molnupiravir trials and two nirmatrelvir/ritonavir trials. In the molnupiravir trials, total viral RNA drops less than non-mutated viral RNA due to PCR detection of drug-mutated viral RNA. Total possible reduction in non-mutated SARS-CoV-2 RNA is less for MOVe-OUT than PLATCOV and PANORAMIC due to higher initial viral loads and lower values of detection in the trials.

These potencies mapped to different reductions in viral load relative to placebo/usual care. Total viral RNA reduction in PANORMIC and PLATCOV exceeded that in MOVe-OUT owing to lower paf **(Fig 6d)**, but also due to a larger viral dynamic range (defined as the distance from baseline viral load to the lower threshold PCR (LOD) **(Fig S10b)**), which allows for a greater observed reduction in viral load. PANORAMIC used LOD = 109 (imputed as 50), and PLATCOV had LOD≅18 (copies/ml), but MOVe-OUT had a higher LOD = 500 copies/ml. PANORAMIC also had much higher average starting viral loads (7.4 log10 (copies/ml)) versus PLATCOV (5.8 log10 copies/ml) and MOVe-OUT (6.8 log10 copies/ml) **(Fig S10b)**. Molnupiravir approached maximal possible total viral RNA reduction in PANORAMIC and PLATCOV, whereas protease inhibitors could still achieve greater viral load reduction at lower paf **(Fig 6d)** as recently observed with ensitrelvir.^27^

The greater possible reduction in total viral load for protease inhibitors relative to mutagenic drugs like molnupiravir owes to different mechanisms of action. Model projected reductions in non-mutated viral RNA reduction in PANORMIC and PLATCOV approximated viral load reductions observed in EPIC-HR and PLATCOV on nirmatrelvir / ritonavir **(Fig 6e)**. This suggests that PCR detection of mutated viruses underestimates true molnupiravir potency, and that a more potent mutagenic agent could accrue further virologic benefit even if the total reduction in viral RNA does not increase.

### Optimization of molnupiravir therapy to avoid viral rebound

Instances of viral rebound were observed in PLATCOV and PANORAMIC and have been observed following molnupiravir treatment.^4, 28^ We analyzed higher doses and prolonged therapy and noted that as with nirmatrelvir, prolonging therapy is a better method to prevent rebound than increasing dose **(Fig S12)**.

## Discussion

We recapitulated the virologic results of three clinical trials for molnupiravir with our combined clinical trial simulation VID-PK-PD models. Model output highlights key differences in viral load reduction between the trials and identifies mechanistic differences to explain this. The MOVe-OUT trial was associated with significantly less reduction in viral load between treatment and control arms than the other two trials. Accordingly, the average drug efficacy over the dosing interval (53%) was lower in this trial, and our estimate for paf was 2.64, signifying marginally lower potency in vivo than in vitro. This result is compatible with 0.4-fold lower median EC50 values for molnupiravir in vitro against omicron relative to prior variants,^26^ though explanations other than viral variant cannot be ruled out. The higher paf in MOVe-OUT permitted drug troughs below the EC50, limiting potency throughout the dosing interval. In PLATCOV and PANORAMIC, our model suggests drug levels remain above the in vivo EC50 throughout the dosing interval though potency does fluctuate according to drug level.

The projected drug efficacy in PLATCOV and PANORAMIC was considerably higher (0.94 and 0.95, respectively), and the paf was estimated to be 0.14 and 0.13, indicating greater potency in vivo than in vitro. We have applied our clinical trial simulation technique to multiple drugs for SARS-CoV-2,^11^ HSV-2,^22, 23^ and HIV,^29^ and this is the first time we have identified this trend.

Allowing molnupiravir to be more potent in vivo in our modeling of PLATCOV and PANORAMIC was necessary to capture the much greater reduction in total viral RNA relative to off-treatment in these studies (1.09 log and 0.94 log respectively versus 0.3 log in MOVe-OUT).

A key outcome of our analysis is the prediction that SARS-CoV-2 PCR likely underestimates molnupiravir potency because it still detects drug-mutated viral RNA.^6^ This appears to be most significant when antiviral potency is higher, as in PANORAMIC and PLATCOV, leading to a 0.48-0.59 log underestimation of reduction in non-mutated virus. Our results suggest that use of standard PCR for assessing SARS-CoV-2 levels may lead to underestimation of drug potency. Multi probe assays as have been used for the HIV reservoir may improve specificity for viruses that remain intact and replication competent.^30^ Viral culture is potentially a useful metric but lacks sufficient sensitivity and precision and is too labor intensive to serve as a viable trial endpoint.^13^ In the ferret model, molnupiravir lowered levels of culturable virus while not lowering total viral RNA levels relative to control.^10^

A further consideration of our analysis is selection of optimal virologic endpoints in clinical trials. The PLATCOV study demonstrates that viral clearance slope is an efficient and robust metric to identify potency after enrollment of a limited number of trial participants.^4^ It is advantageous relative to viral load reduction as it incorporates all viral load data points in the analysis. We observed small non-statistically significant differences between clearance slopes of total versus non-mutated viral RNA in simulations of all three trials. However, we observed significant reductions in viral area under the curve during therapy in all three simulated trials, suggesting that this may be an even more sensitive trial endpoint. In viral dynamic models, viral AUC maps directly to surface area of total infected cells providing a mechanistic underpinning of why this may be a useful endpoint.^31, 32^

Our results suggest that viral loads may vary according to drug level given molnupiravir’s short plasma half-life. A sub-study within future trials comparing viral loads between drug trough and peak would be useful for the field. This could validate our model’s prediction that even small reductions in viral RNA may be associated with substantial reductions in total viral AUC particularly with a short half-life drug. Even minor reductions in viral load may be associated with substantial clinical benefit in this case. These fluctuations may be less evident if the intra-cellular half-life of the drug is longer or if PK measures in our model underestimate true drug levels in PANORMIC due to older age or impaired renal clearance. Model accuracy would be improved if viral load and PK data were available from the same patients in similar trials.

Another key practical outcome is that as for nirmatrelvir, extension of molnupiravir therapy to 10 days is likely to prevent rebound, though our simulations do not suggest any benefit from increasing dose.^11^ This suggests that prolonging therapy or using longer half-life agents is ideal for treating SARS-CoV-2.^33^

Each trial represented a unique set of issues for model fitting. In MOVe-OUT, because the mean viral kinetics curve of treatment arm differed only slightly from that of the control arm, the model without drug provided reasonable fit to the treatment arm. Nevertheless, the paf was identifiable for this trial indicating that the model was able to detect and specify the very limited potency of the drug. The fact that the drug’s potency and clinical efficacy appears to have increased with introduction of the omicron variant demonstrates a massive challenge for the therapeutics field: as with vaccines, trials performed when prior variants were circulating may prove less relevant as new variants continually emerge. A priority should be retesting existing agents against newly emerging variants in small nimble trials such as PLATCOV, with viral load endpoints.

For PLATCOV, the model for the treatment arm matched the trial data precisely and identified the paf. The drug achieved nearly maximal observed viral load reduction in this trial. We identified a similar trend for PANORAMIC. It is notable that the model had the flexibility to account for different viral loads between these trials by predicting more rapid innate immune responses in PLATCOV.

We arrived at similar estimated paf in the PLATCOV and PANORAMIC trials, which agreed when using two separate methods: fitting to individual viral loads and fitting to mean viral load reduction trial endpoints. This suggests that our approach using in silico cohorts and fitting to population level outcomes produces reliable results.^11^ In many cases, it remains challenging for academic researchers to obtain individualized data from industry sponsored trials. Therefore, it is important that the endpoint fitting approach be considered when this is the only data available.

Finally, our results highlight challenges in trial design associated with the selection of PCR assays and the corresponding limits of detection. Each trial reported results with a different limit of detection which in turn impacts the degree of viral load decrease that can be observed. Initial viral loads were notably higher in PANORAMIC and MOVe-OUT than PLATCOV. The equivalent viral loads between PANORAMIC and PLATCOV may reflect a more sensitive PCR in the PANORAMIC study as past immunity has consistently predicted lower viral loads and more rapid viral elimination for omicron variants.^12, 34^ On the other hand, PANORMIC viral loads could have been higher than in PLATCOV despite both enrolling omicron infections due to an older and less healthy population, Ideally, equivalent internationally standardized PCR quantitation would be used across all trials.

Our study has a few limitations. The estimated paf is based on the in vitro assay data against delta variant in Calu-3 cells. In vitro EC50 is sensitive to assay conditions, including cell type, the variant of concern, the multiplicity of infection (MOI), and specific lab. In general, the inability of in vitro assays to match in vivo conditions makes it an unreliable proxy for the in vivo potency of the drug and explains the necessity of incorporating the paf parameter when simulating an antiviral clinical trial.

In our model, we assumed all mutated viruses are noninfectious. While most drug-induced random mutations reduce the fitness/viability of the virus, the molnupiravir mutation signature has been detected in circulating variants, especially in regions where the drug has been used most commonly.^7^ Furthermore, in the PANORAMIC study, no statistical difference was observed between the rate of culturable samples in the treatment and control arms which implies some mutated viruses might be transmissible.^35^ We justify this assumption in our definition of drug efficacy. The incomplete efficacy in all three trials may reflect subtherapeutic drug levels but could also indicate that not all drug-induced mutations not lower viral infectivity.

PK parameters were estimated using the data from the plasma concentration of healthy individuals with the age range of 19-60 (mean 39.6) years old. The clearance rate of renally cleared drugs often increase with age.^36^ This implies that the paf may be larger in an older population, such as in PANORAMIC participants. Further, we used the plasma concentration in the PD model to calculate the drug efficacy. However, using the drug’s intracellular concentration with a longer half-life, represented by the central compartment of the PK model, could also likely lead to a larger estimated paf.

In summary, we further demonstrate the utility of clinical trial simulation using models that capture drug PK and PD, as well as infection dynamics. In the case of molnupiravir, our results suggest that final viral endpoints should be adjusted based on the drug’s mechanism of action.

## Materials and Methods

### Study Design

We developed a viral dynamics model recapitulating viral load data collected from symptomatic individuals in the NBA (National Basketball Association) cohort.^12^ We used a two-compartment model to reproduce the PK data of molnupiravir.^19^ For clinical trial simulation, we constructed a virtual cohort by randomly selecting 400 individuals from the NBA cohort, matching the trial populations regarding vaccine status and history of infection as closely as possible given cohort characteristics. Due to lack of individual PK data, we used estimated population PK parameters for all individuals in the virtual cohort. The PD parameters for each individual were randomly selected from a log-normal distribution with their means and standard errors estimated based on in vitro assay data. We fit the combined viral dynamic and PKPD model to the average change in viral load from the baseline of the control and treatment arms of three previously published molnupiravir clinical trials.^2-4, 37^ Comparing our model to the control arms validated our viral dynamic model and demonstrated how well our virtual cohorts represent the trial control arms. We used the average data from the treatment arms to estimate the potency adjustment factor (paf) by maximizing the R^2^ of the fit.^11^ We also fit to individual viral load trajectories in PLATCOV and PANORAMIC using the mixed-effect population approach implemented in Monolix^38-40^ and obtained both individual paf values and a population distribution.

### Viral load data

The NBA cohort dataset published by Hay et al consists of 2875 documented SARS-CoV-2 infections in 2678 people detected through frequent PCR testing regardless of symptoms.^17^ We used the viral load data from 1510 infections in 1440 individuals that had at least 4 positive quantitative samples to estimate viral dynamic parameters. We used parameter sets estimated for the symptomatic subpopulation of these individuals to construct virtual cohorts.^12^

### Clinical trial data

We used viral load data from three molnupiravir clinical trials. MOVe-OUT by Jayk Bernal et al. included 544 and 549 symptomatic high-risk individuals in the control and treatment arms, respectively.^2^ We obtained the average change in viral load data of the control and treatment arms as shared in the supplementary material of their published manuscript in Table S6. Nasal viral load was measured using PCR assay on days 0, 3, 5, 10, and 14 after the treatment start day and adjusted by the baseline viral load. A lower limit of detection (LOD) of 500 copies/ml was used in this trial. The treatment was started within five days of symptom onset.

PLATCOV by Schilling et al. is an open-label, randomized, controlled adaptive trial with 85 and 58 symptomatic, young, healthy individuals in the control and molnupiravir treatment arms, respectively.^4^ The oropharyngeal samples from each participant were collected daily on days 0 through 7 and on day 14 after the treatment start day, and viral load was measured using PCR assay. We used the individual viral load data published by the authors for model fitting. From PLATCOV, we averaged over the two oral samples collected from each individual for individual-level viral load fitting, and averaged over all samples collected then calculated viral load drop from baseline for the trial endpoint fitting approach. In both cases, when comparing simulation results with data we used the maximum LOD reported in the published data (∼1.26 log).

PANORAMIC is a platform adaptive randomized, controlled trial with 42 and 38 symptomatic, vaccinated individuals with at least one risk factor in the control and treatment arms, respectively.^3^ Nasal viral load was measured using PCR and samples were collected on days 0 through 6 and on day 13. We used individual viral load data shared by the authors and adjusted by baseline viral load to obtain mean viral load drop from the baseline. Mean days since symptom onset at baseline (sd) were 2.4 (0.78) for the treatment arm and 2.5(1.12) for the control arm. The lower limit of detection of 100 copies/ml (imputed as 50 copies/ml) was used.

In all three trials, the study participants were treated with 800mg molnupiravir twice per day, for five days.

### PK data

Mean plasma concentration data of molnupiravir were obtained by digitizing Figure 3 of the phase I trial by Painter et al. using WebPlotDigitizer.^19^ The data used in this paper belong to the multiple-ascending part of the phase I trial where six participants were given 50, 100, 200, 300, 400, 600, and 800 mg of molnupiravir twice daily for 5.5 days, and their plasma concentrations were measured after the first and last doses.

### PD data

The data on drug efficacy was obtained from experiments performed at the University of Washington. The efficacy of molnupiravir was measured against the delta variant of SARS-CoV2 in Calu-3 cells (human lung epithelial). Briefly, Calu 3 cells were treated with varying concentrations of molnupiravir prior to infection with SARS-CoV-2 (delta isolate) at a multiplicity of infection of 0.01. Antiviral efficacy and cell viability (of non-infected cells treated with drugs) were assessed as described.^9, 11^ There were five replicates per condition, pooled from 2 independent technical experimental repeats (one experiment with triplicate conditions, one experiment in duplicate conditions).

### Viral dynamics model

We used data-validated model of SARS-CoV-2 dynamics to model the viral load of symptomatic individuals with SARS-CoV-2 infection.^12^ This model assumes that susceptible cells (S) are infected at rate βVS by SARS-CoV-2 virions. The infected cells go through a non-productive eclipse phase (I_E_) before producing viruses and transition to becoming productively infected (I_P_) at rate *κ*I_E_. When encountering productively infected cells, the susceptible cells become refractory to infection (R) at the rate *ϕ*I_P_S. Refractory cells revert to a susceptible state at rate *ρ*R. The productively infected cells produce virus at the rate πI_P_ and are cleared at rate *δ*I representing cytolysis and the innate immune response that lacks memory and is proportional to the amount of ongoing infection. If the infection persists longer than time τ, then cytotoxic acquired immunity is activated, which is represented in our model by the rate *m*I_P_. Finally, free virions are cleared at the rate *γ*. Of note, this model, previously proposed by Ke et al. was selected against other models based on superior fit to data and parsimony.^41^ The model written as a set of differential equations has the form,

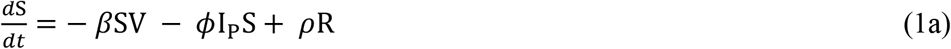

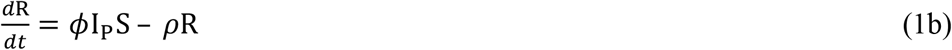

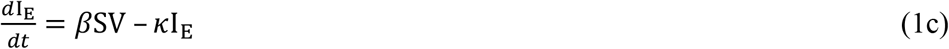

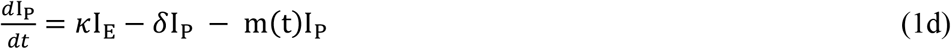

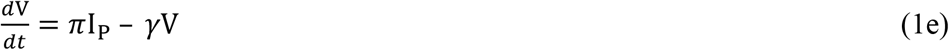

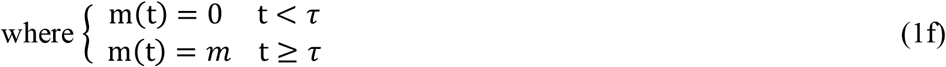

To estimate parameter values, we fit the model to viral load data from the NBA cohort using a mixed-effect population approach implemented in Monolix. Details on the model selection and fitting process can be found in Owens et al.^12^

We start the simulations with 10^7^ susceptible cells. The initial value of the refractory cells is assumed to be zero since the interferon signaling is not active prior to infection. We further assume there are no infected cells (eclipse or productive) at the beginning of the infection. We fix the level of inoculum (V_0_) at 97 copies/ml for each individual.

To resolve identifiability issues, we also fixed two parameter values, setting the inverse of the eclipse phase duration to *κ* = 4, and the rate of clearance of virions to *γ* = 15.^12^

### PK model

We used a two-compartmental PK model which includes the amount of drug in the GI tract (A_GI_), the plasma compartment (A_P_), and the respiratory tract (A_L_). The drug is administered orally, passes through the GI tract and gets absorbed into the blood at the rate *κ*_*a*_. The drug then transfers from the blood into the peripheral compartment (or the respiratory tract) at the rate *k*_*PL*_. The metabolized drug transfers back into the plasma at the rate *κ*_*LP*_ from where it clears from the body at the rate *k*_*CL*_. The model in the form of ordinary differential equations is written as,

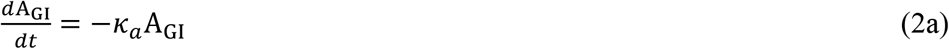

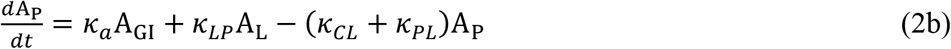

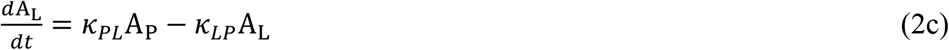

We used Monolix and a mixed-effect population approach to estimate the parameters and their standard deviations. With the initial condition of (A_GI_ = DoSE, A_P_ = 0, A_L_ = 0); we fit 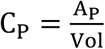 to the plasma concentration data where Vol is the estimated plasma volume. Details on parameter values and the error model provided in **Table S1**.

### PD model

For the pharmacodynamics model we used Hill equation, 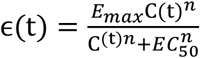, where C(t) is the drug concentration in plasma, *E*_*m*a*x*_ is the maximum efficacy, *n* is the Hill coefficient, and *EC*_50_ is the drug concentration in plasma required for 50% efficacy. We used least-squared fitting to obtain the three parameters (*E*_*m*a*x*_, *n*, and *EC*_50_) and their standard deviations. The average drug efficacy over the treatment period is calculated using the formula,

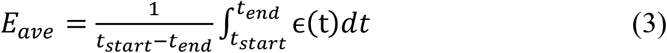

Where t_start_ and t_end_ are the treatment start day and end day, respectively.

### Combined PKPD and VL models

The plasma concentration of molnupiravir obtained from the PK model is used in the PD model to obtain time-dependent efficacy. Since molnupiravir imposes lethal mutations during the viral replication process, in our model, a portion of all viruses produced by an infected cell, measured by ϵ(t), are mutated (*V*_*m*_) and therefore assumed to be non-infectious, with the addition that most detected viral RNA pre-treatment is also non-infectious. The production rate of non-mutated viruses is decreased by a factor of (1 ™ ϵ(t)). Equation 1e is written as,

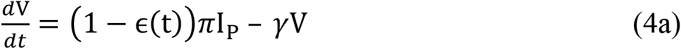

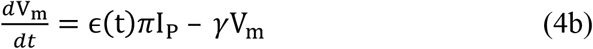

Total viral load (*V* + *V*_*m*_) is used to fit the PCR assay data from each trial.

### Fitting the combined model to individual viral load data in the PLATCOV trial

We used the population mixed-effect approach implemented in Monolix to estimate each individual’s viral dynamics parameters and the potency adjustment factor (paf). Due to the lack of data from the initial phase of infection in the PLATCOV and PANORAMIC trials, we include the data from Omicron-infected individuals in the NBA cohort in the fitting populationto inform the model about the initial phase of infection. We fixed the PK parameters to the estimated population values (**Table S1**), and the PD parameters other than EC50 to the in vitro estimated population values (**Table S2**). We used the study category (NBA vs PLATCOV and PANORAMIC) as a covariate for t_0_ (timing of infection) and τ (timing of the adaptive immune response) since the first recorded positive test is likely much later for the clinical trials. In the NBA study, samples were collected almost daily regardless of symptoms often leading to pre-symptomatic detection, while in the two clinical trials, the baseline measurement occurred after symptom onset, trial enrollment and consent.

### Construction of a virtual cohort

To generate a cohort for our simulated clinical trials, we randomly selected 400 individuals (for each arm) from the unvaccinated symptomatic subpopulation of the NBA cohort for MOVe-OUT and the vaccinated, Omicron infected subpopulation for PLATCOV and PANORAMIC and used their individual viral load parameters estimated by fitting our viral dynamics model to the data. A sample size of n=400 (out of 822 vaccinated individuals with Omicron infection) was used to mimic a large-scale clinical trial and maintain relatively low overlap between virtual cohorts used in each arm of the simulations and between different simulations. Since the time of symptom onset is not available for all individuals in the NBA data, we randomly drew an incubation period for each individual from gamma distributions with variant-specific parameters estimated by Gamiche et al.^42^ The start of treatment relative to symptom onset was randomly selected according to a uniform distribution for MOVe-OUT and PLATCOV, and a logit normal distribution for PANORAMIC with limits of [0,5] days and mean and standard deviation reported in the PANORAMIC trial for control and treatment arms. The same population PK parameters were assigned to each individual. The relevant dose in each scenario was added to the A_GI_ compartment (the absorption equation) of the PK model (**eq 2a**) at each dosing timepoint (t=0, 0.5, 1, 1.5, …., 4.5 days). For each dose, the appropriate PK parameter were used **(Table S1)**. PD parameters were also randomly drawn from a log-normal distribution with the estimated mean and standard deviation. The standard deviation of the PD parameters represents the accuracy of the assays and not individual variability.

### Potency adjustment factor (paf)

The paf is defined as,

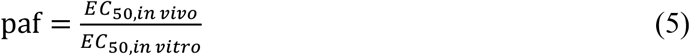

We estimated the paf by maximizing *R*^*2*^ when fitting the change in viral load of the treatment arm of our simulation to the change in viral load observed in the treatment arm of the clinical trial on day 0 through day 7.

### Measuring rebound probability

A viral load rebound in the treatment arm was defined when the viral load at any time after treatment ended exceeded the viral load at the end of the treatment by ≥1 on a log_10_ scale. In the control group, viral rebound was defined in patients who had at least two peaks with minimum height of 1000 copies/ml in their viral load trajectories and the second peak was ≥ 1 log higher than its preceding local minimum.

### Software

All the model fittings in this study were performed using Monolix version 2023R1.

The data analysis and simulations were performed using Python 3.9.12.

## Data availability

The data analyzed in this work was previously published by Hay et al., Schilling et al. and Standing et al., and is available at:

https://github.com/gradlab/SC2-kinetics-immune-history and https://github.com/jwatowatson/PLATCOV-Molnupiravir/tree/V1.0

https://zenodo.org/records/10375295

Pharmacodynamics data is available on GitHub at https://github.com/sEsmaeili/MolnupiravirModeling

## Code availability

All codes and materials used in the analysis are available on GitHub https://github.com/sEsmaeili/MolnupiravirModeling

## Author Contributions

Conceptualization: JTS, SE, KO

Methodology: JTS, SE, KO

Software: SE, KO

Investigation: JTS, SE, SJP, JW

Formal analysis: SE

Writing – original draft: JTS, SE

Writing – review & editing: JTS, SE, KO, SJP, JFS, DML, SZ, JAW, WHKS,

