## Supplementary material for "Molnupiravir clinical trial simulation suggests that polymerase chain reaction underestimates antiviral potency against SARS-CoV-2": Supplmentary material

For

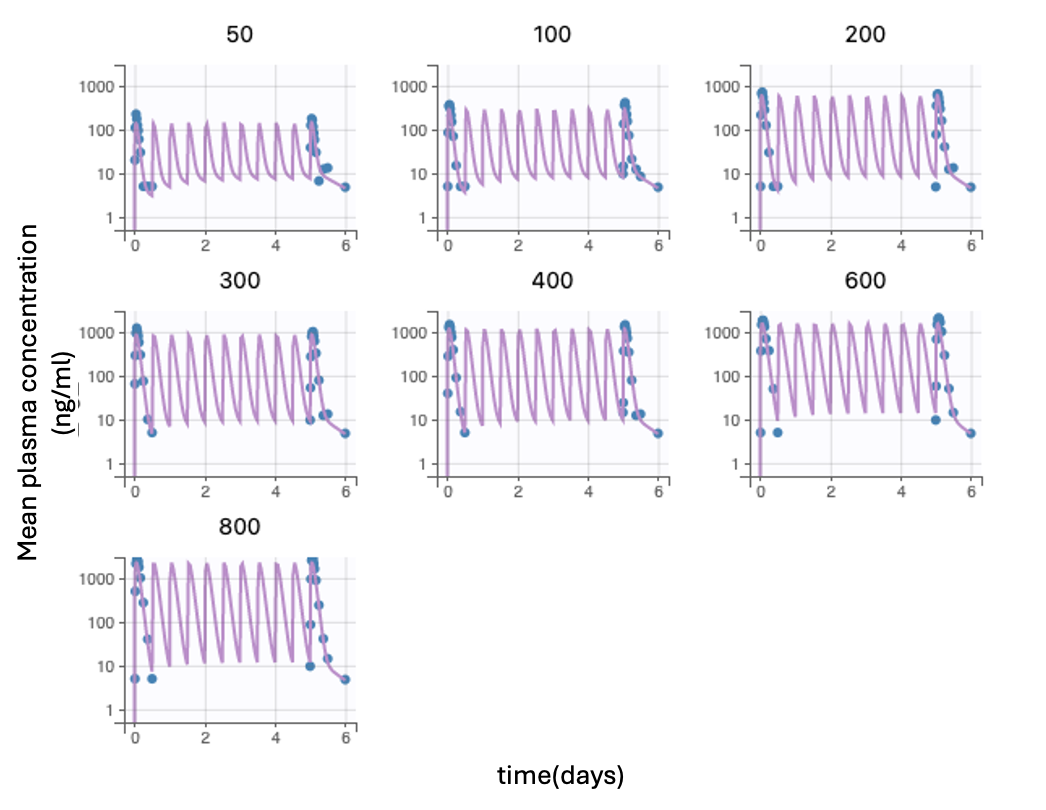

**Figure S1.** **Pharmacokinetics model fit to data.** Datapoints reflect mean plasma concentration of 50, 100, 200, 300, 400, 600, and 800 mg of molnupiravir given twice daily for 5.5 days.

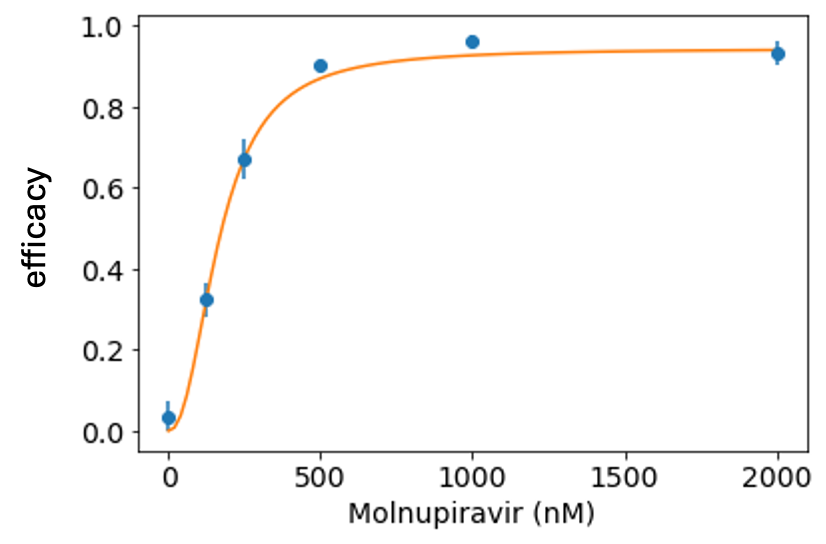

**Figure S2. Pharmacodynamic model fit to in-vitro assay data.** The estimated EC50 is 177 nMOL.

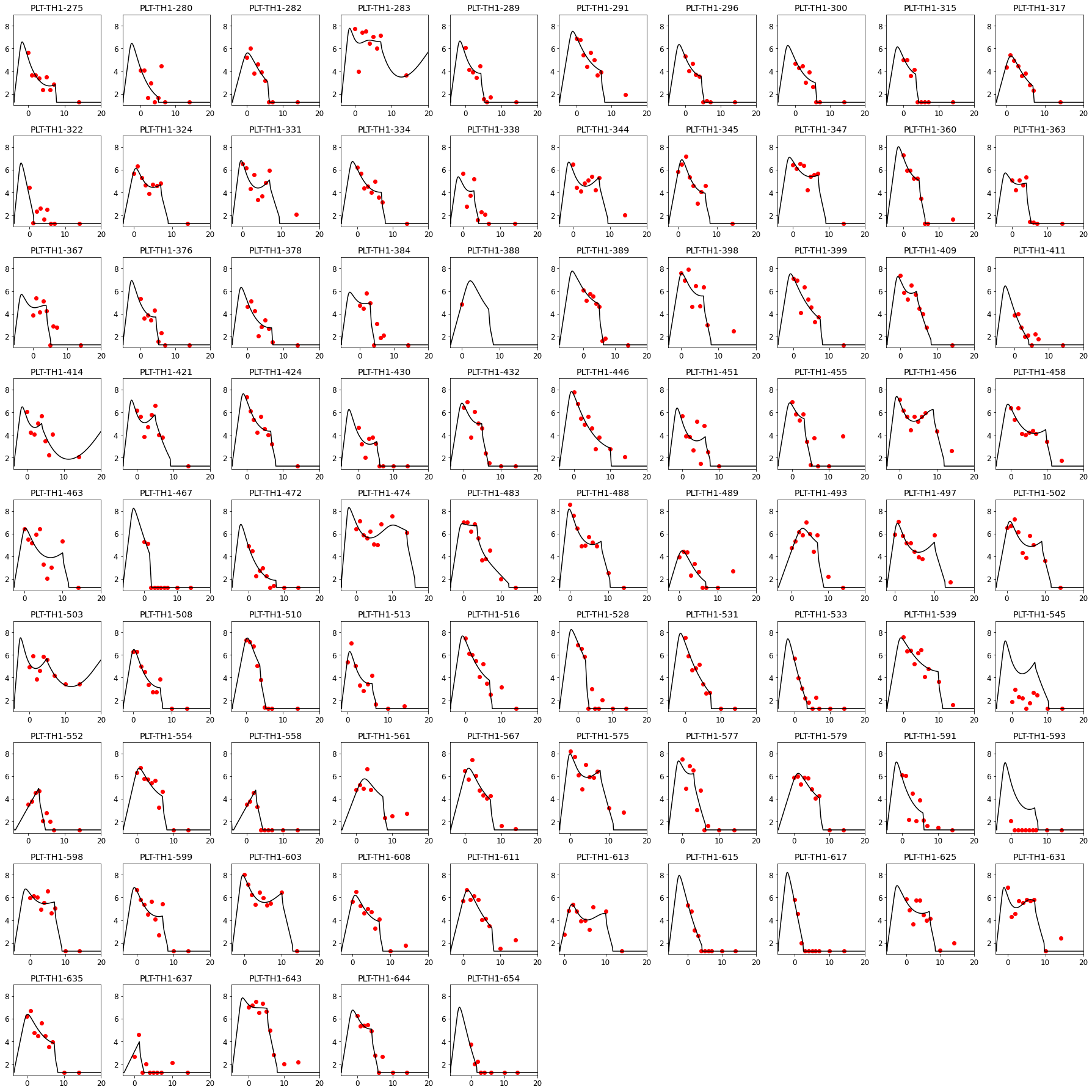

**Figure S3. Individual model fits to viral load data from the control arm of PLATCOV trial.**

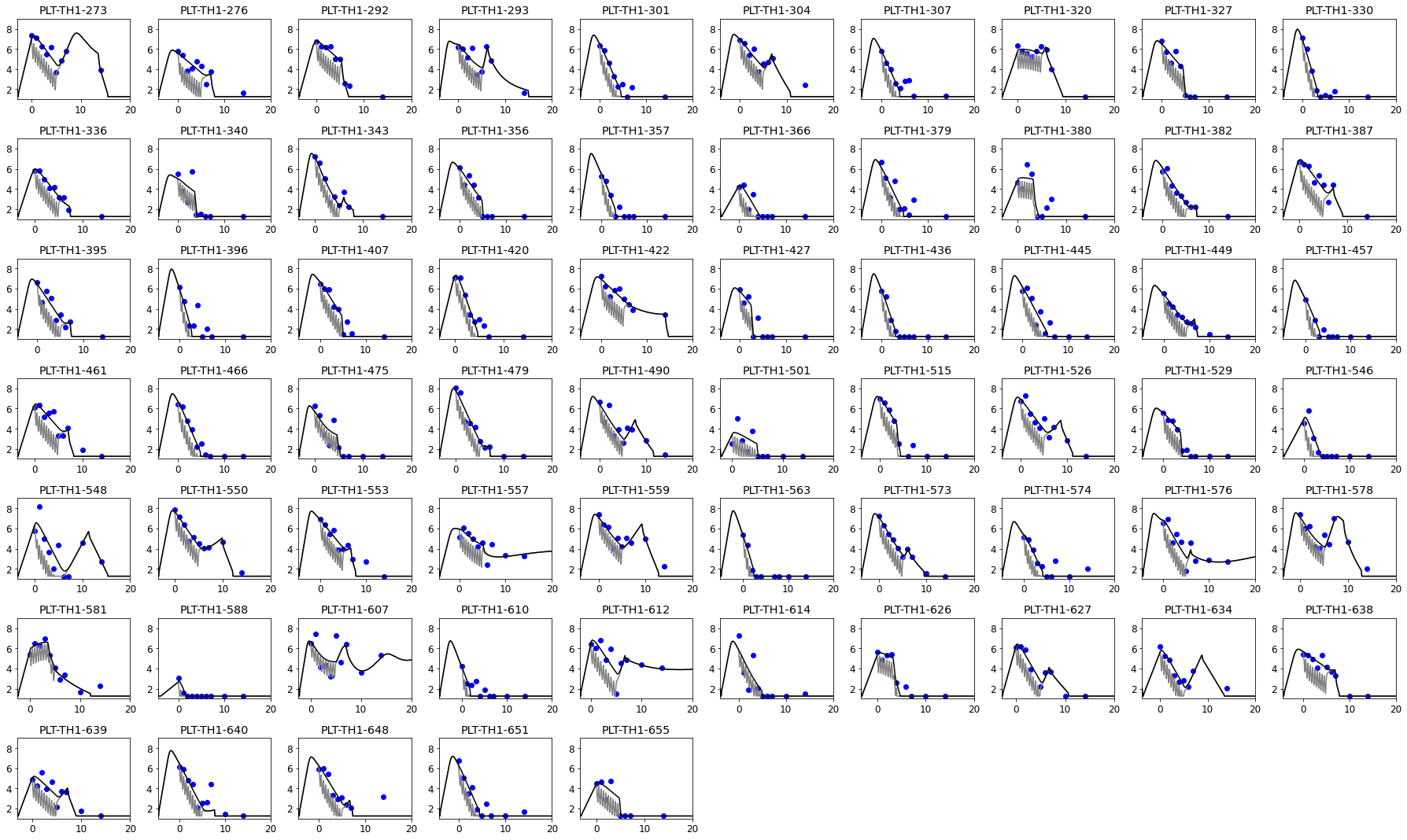

**Figure S4. Individual model fits to viral load data from the molnupiravir treatment arm of the PLATCOV trial.**

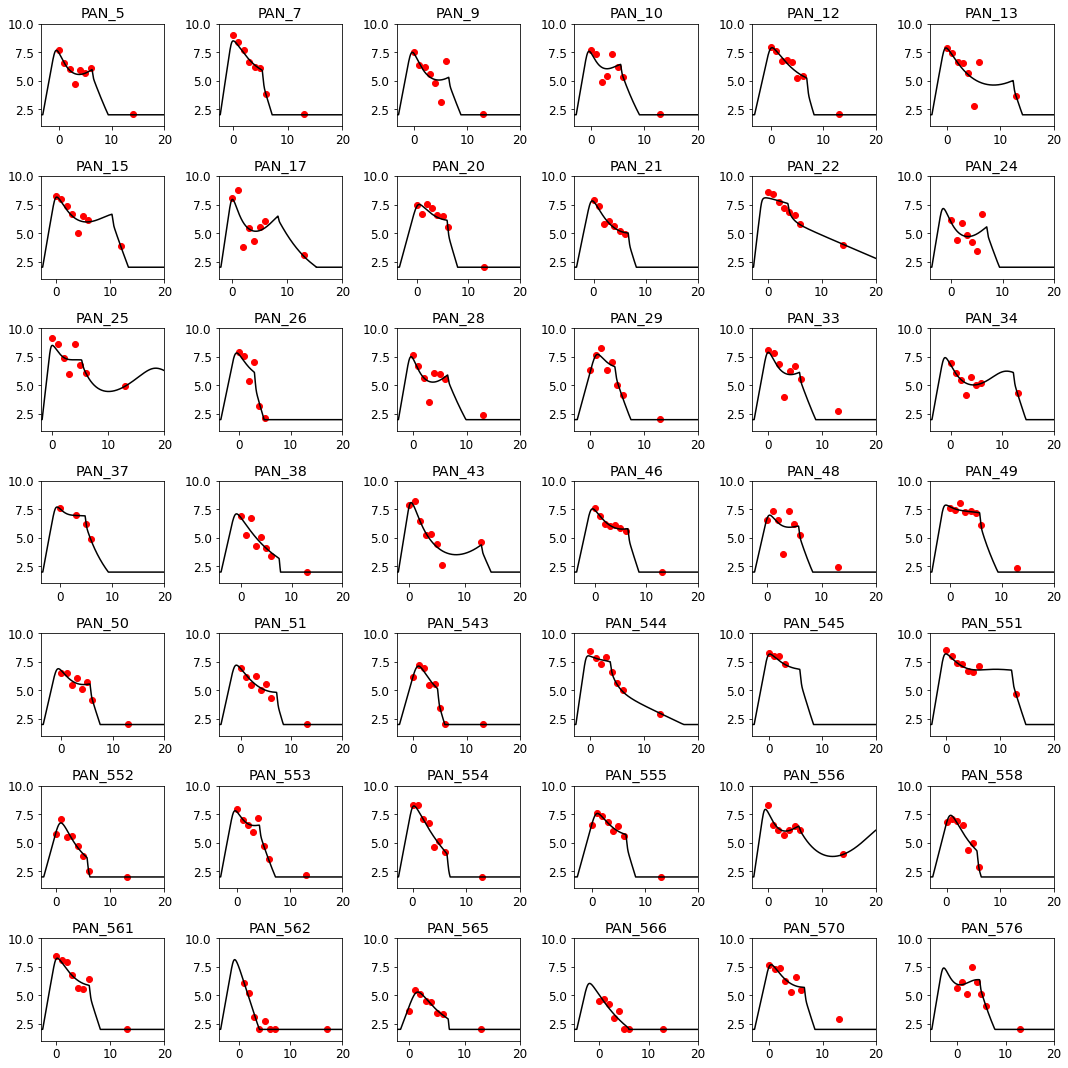

**Figure S5. Individual model fits to viral load data from the control arm of PANORAMIC trial.**

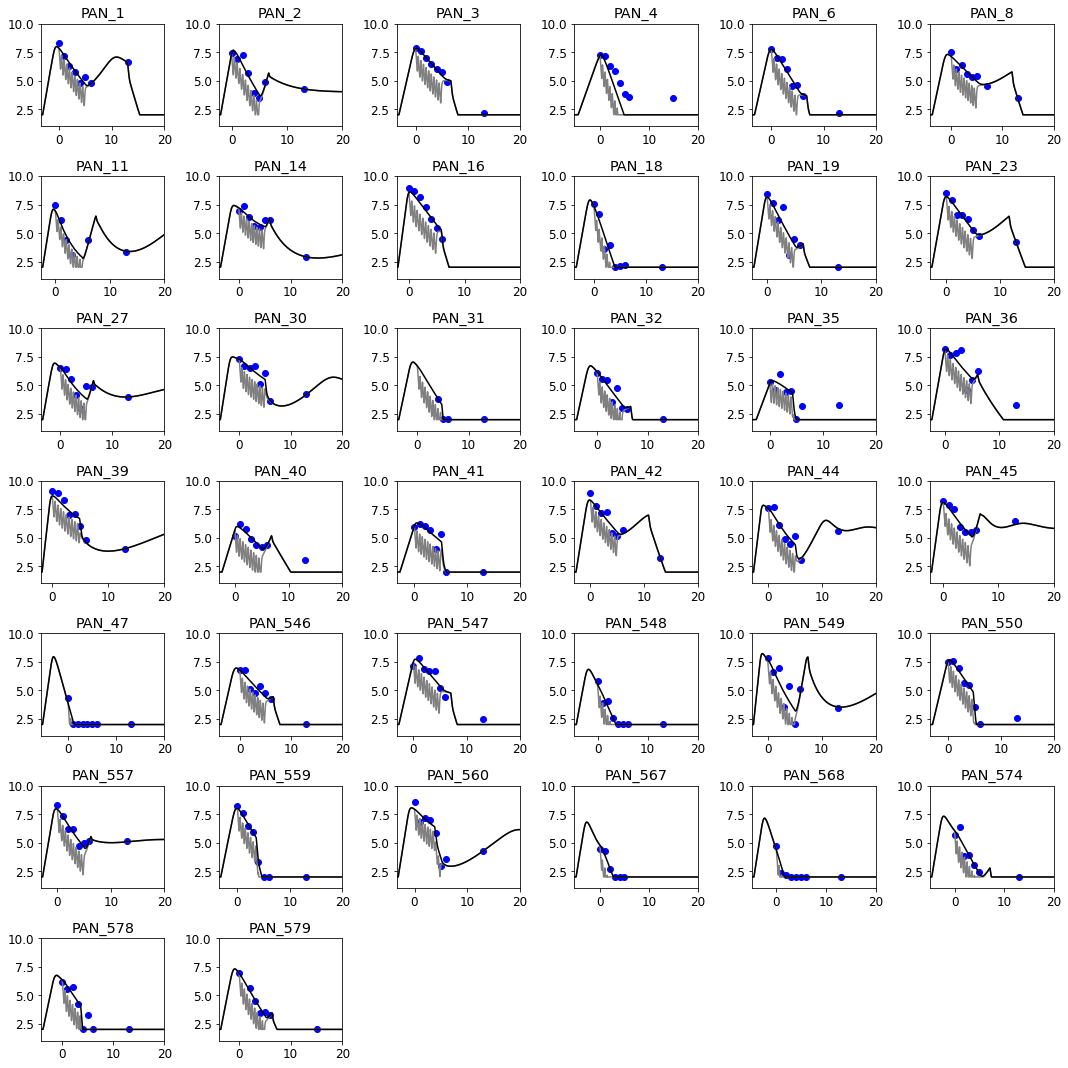

**Figure S6. Individual model fits to viral load data from the treatment arm of PANORAMIC trial.**

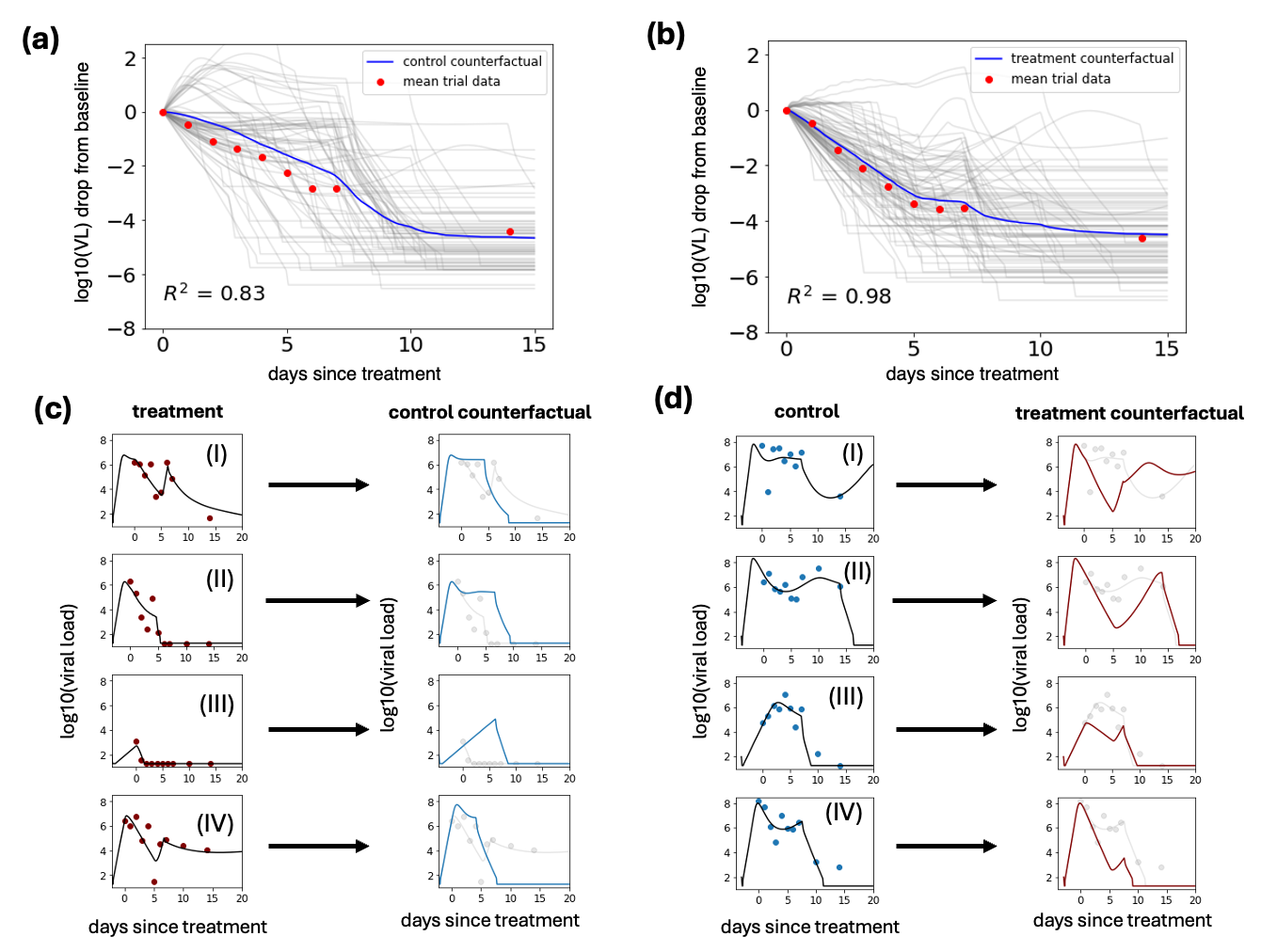

**FigS7. Counterfactual simulations of the control and treatment arms of the PLATCOV trial recapitulate trial outcomes.** (a-b) individual simulated control counterfactual of the treatment arm (a) and the treatment counterfactual of the control arm (b) (in grey) and the mean viral load drop from the baseline (in blue) and the trial data. (c-d) sample model fits to the treatment arm and its control counterfactual simulation (c) and control arm and its treatment counterfactual simulation(d).

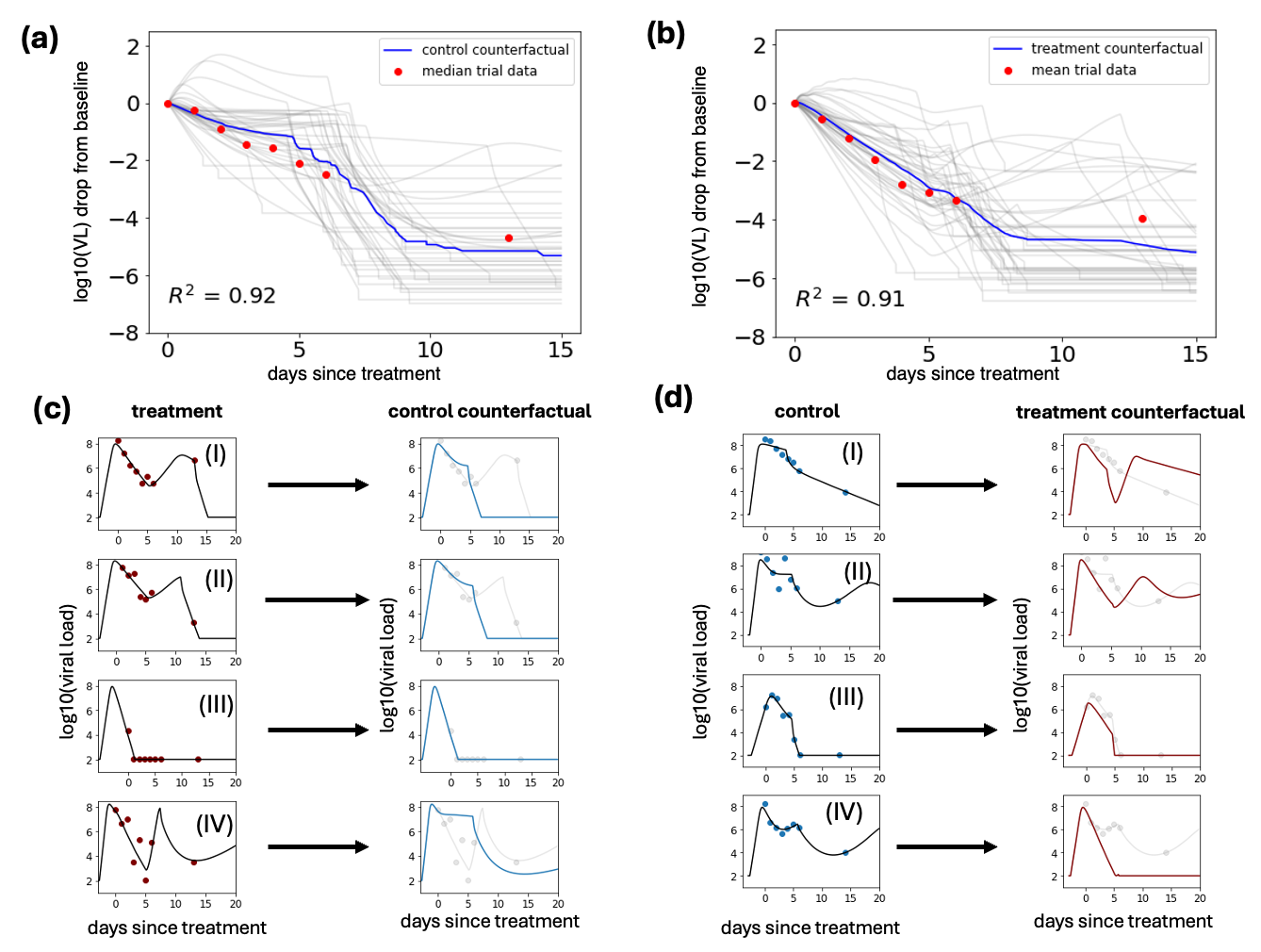

**FigS8. Counterfactual simulations of the control and treatment arms of the PANORAMIC trial recapitulate trial outcomes.** (a-b) individual simulated control counterfactual of the treatment arm (a) and the treatment counterfactual of the control arm (b) (in grey) and the mean viral load drop from the baseline (in blue) and the trial data. (c-d) sample model fits to the treatment arm and its control counterfactual simulation (c) and control arm and its treatment counterfactual simulation(d).

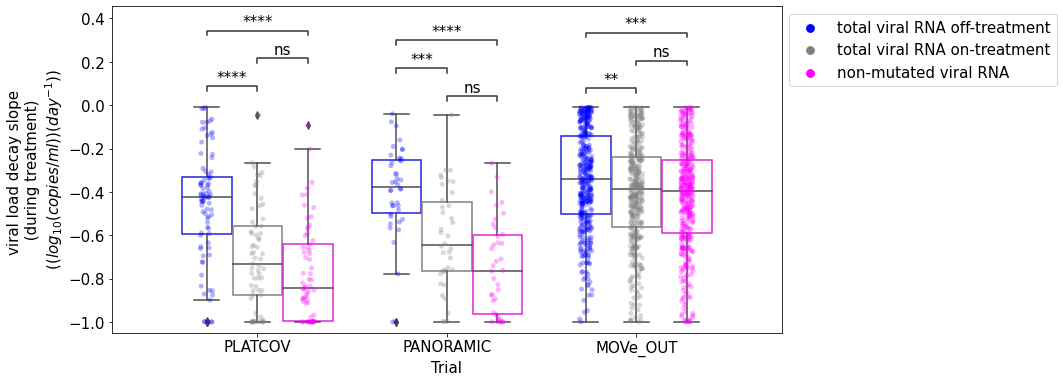

**Figure S9. Comparing the slope of viral decay of total viral RNA on and off-treatment and non-mutated viral RNA in three trials.** There is no significant difference between the rate of decay of total viral RNA on-treatment and non-mutated viral RNA.

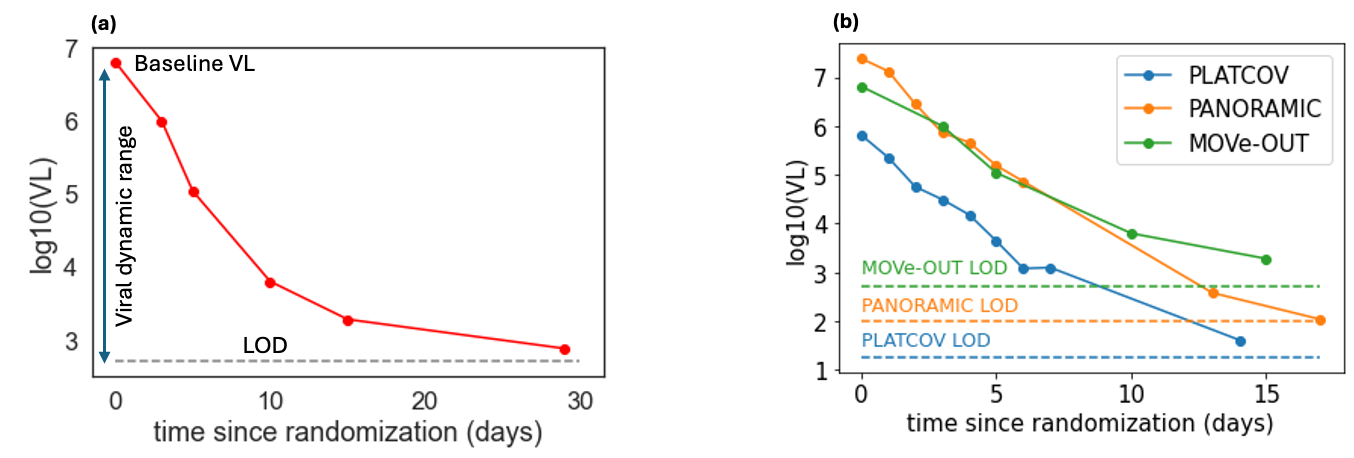

**Figure S10. Different viral dynamic ranges in the three trials.** (a) viral dynamic range is defined as the distance from the baseline to the limit of detection (LOD). (b) comparing the viral dynamic range of the three trials. The dashed lines mark the LOD of each trial.

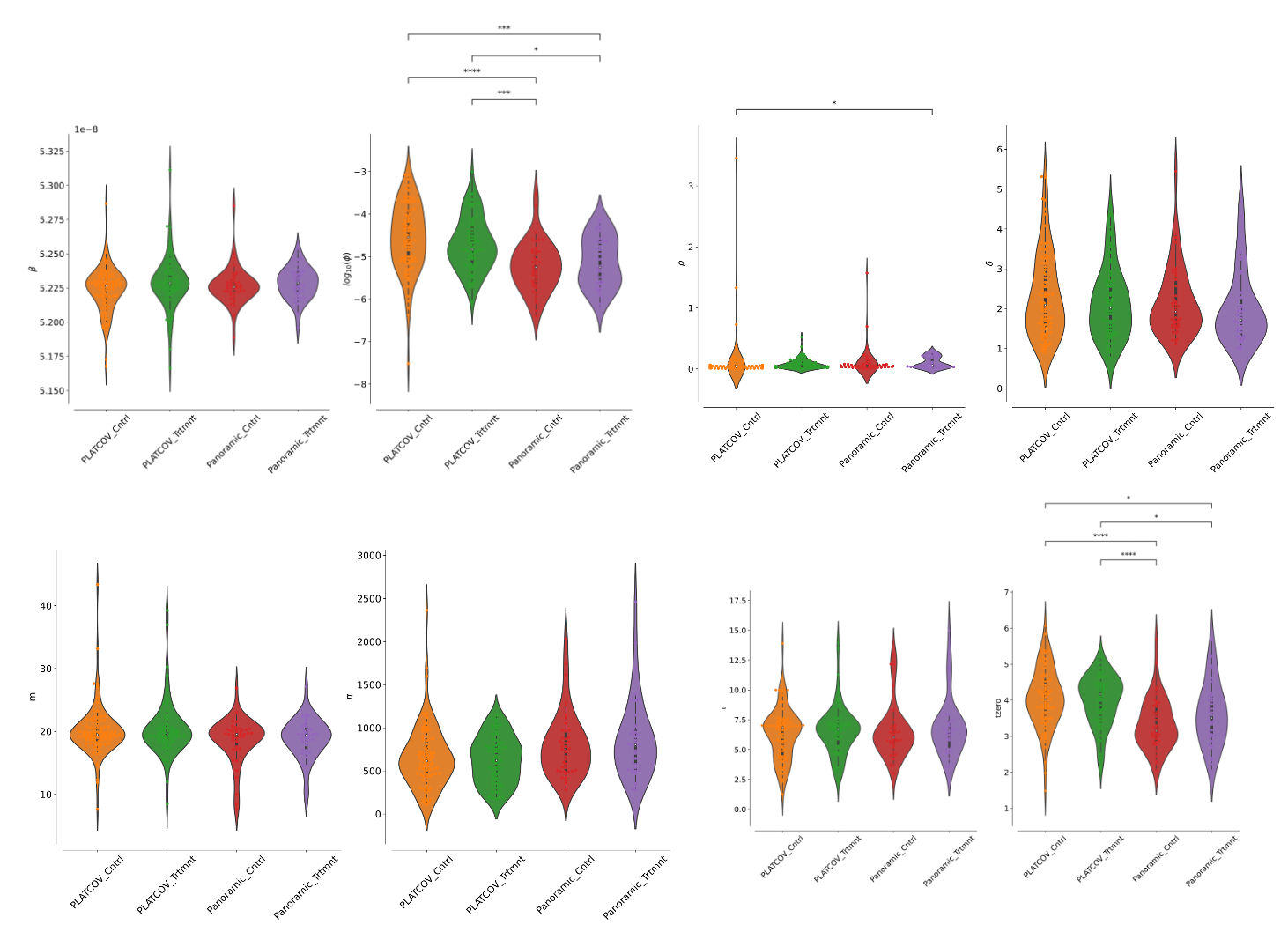

**FigS11. Population parameter distributions two arms of the PLATCOV and PANORAMIC trials**. p-values were obtained by performing two-sided Mann-Whitney U-test (∗: 0.01 < 𝑝 ≤ 0.05,∗∗: 0.001 < 𝑝 ≤ 0.01,∗∗∗: 0.0001 < 𝑝 ≤ 0.001,∗∗∗∗: 0.00001 < 𝑝 ≤ 0.0001).

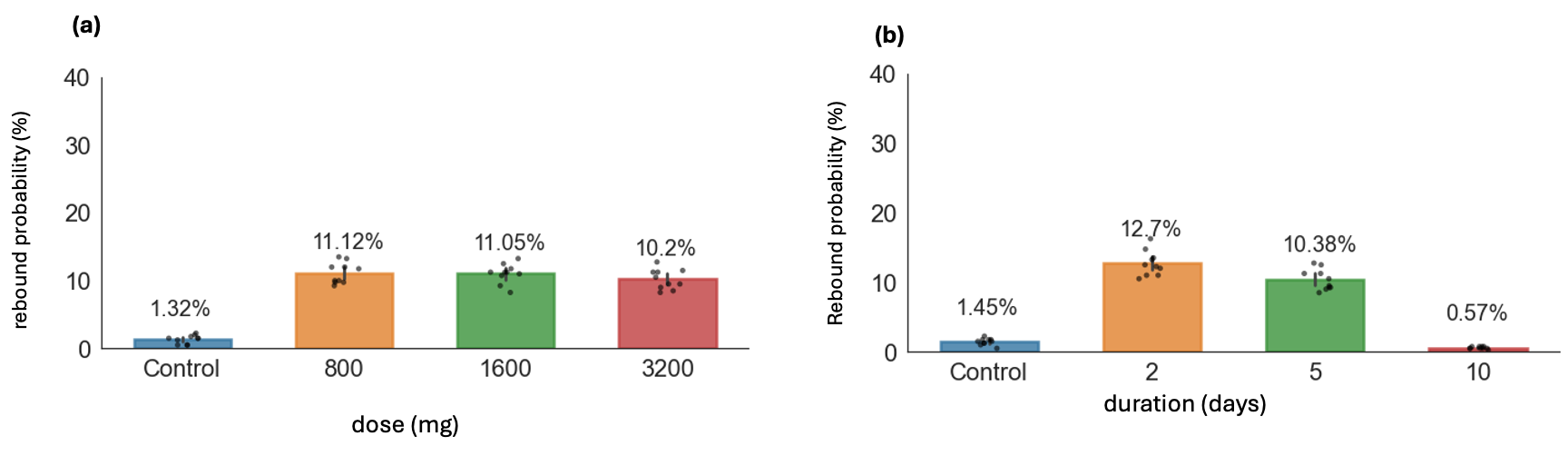

**Figure S12. Longer treatment limits the chance of rebound.** (a) rebound probability for different doses of molnupiravir administered twice daily for 5 days. (b) rebound probability following 800mg of molnupiravir given twice daily for different durations.

| **Dose (mg)** | $\boldsymbol{\kappa}_{\boldsymbol{a}}$(day^-1^) | $\boldsymbol{\kappa}_{\boldsymbol{LP}}$(day^-1^) | $\boldsymbol{\kappa}_{\boldsymbol{CL}}$(day^-1^) | $\boldsymbol{\kappa}_{\boldsymbol{PL,1}}\boldsymbol{(}mg^{-1}\boldsymbol{)}$ | **Vol** (ml) | $\boldsymbol{\alpha}$ |
| --- | --- | --- | --- | --- | --- | --- |
| **50** | 24.38 | 1.32 | 18.98 | 330.31 | 133637.08 | -1.01 |
| **100** | 23.87 | 1.31 | 19.09 | 325.60 | 134136.89 | -1.01 |
| **200** | 24.41 | 1.3 | 19.18 | 325.70 | 134947.57 | -1.01 |
| **300** | 21.79 | 1.3 | 19.06 | 324.84 | 133957.75 | -1.01 |
| **400** | 22.75 | 1.3 | 19.07 | 326.64 | 134158.16 | -1.01 |
| **600** | 16.71 | 1.31 | 18.91 | 323.64 | 133107.91 | -1.01 |
| **800** | 20.16 | 1.31 | 19.03 | 324.14 | 133791.45 | -1.01 |

**Table S1. Estimated PK parameters.** Parameters were estimated by fitting PK model to mean plasma concentration. The transition rate between the plasma and peripheral compartment was dose dependent following a powerlaw relationship ($\kappa_{PL}=\kappa_{PL,1}*Dose^{\alpha})$. The parameters were estimated using mixed-effect population approach in monolix.

| Parameter (unit) | Symbol | Mean | Standard Error |
| --- | --- | --- | --- |
| Maximum efficacy (%) | $E_{max}$ | 1 | NA |
| Drug concentration to provide 50% efficacy ($nMol$ ) | EC_50_ | 177 | 6.6 |
| Hill coefficient | $n$ | 2.05 | 0.22 |

**Table S2.** Estimated PD parameters by fitting to in-vitro assay data, using least square method.

| **Trial** | Clearance half lives in hours: median [IQR] | | |
| --- | --- | --- | --- |
|  | **total viral RNA off-treatment** | **Total viral RNA on-treatment** | **Non-mutated viral RNA** |
| **PLATCOV** | 17.03, [12.19, 21.86] | 9.88, [8.24, 12.95] | 8.56, [7.26, 11.25] |
| **PANORAMIC** | 19.19, [14.51, 28.42] | 11.21, [9.42, 16.23] | 9.45, [7.49,12.07] |
| **MOVe-OUT**  **(virtual cohort)** | 21.27, [14.33, 50.57] | 18.58, [12.89, 30.44] | 18.22, [12.27, 28.64] |

**Table S3.** Simulated viral clearance half-life for each trial for total viral RNA on and off treatment and non-mutated viral RNA on treatment. Median clearance half-life for total viral RNA off treatment and on treatment in PLATCOV 15.5 and 11.6 hours respectively, within the IQR range and within 15 hours of the model estimates for PLATCOV.

| Parameter (unit) | Symbol | Population Mean | Standard error | Std dev. of random effects | Distribution | Source |
| --- | --- | --- | --- | --- | --- | --- |
| viral infectivity  (log_10_ (RNAcopies/mL)^-1^ day^-1^) | log_10_$\beta$ | -7.28 | 1.9e-3 | 1.8e-2 | normal | estimated |
| viral production rate  (log_10_ day^-1^ ) | log_10_$\pi$ | 2.76 | 1.12e-2 | 0.32 | normal | estimated |
| rate at which refractory cells revert to susceptible state  (log_10_ day^-1^ ) | log_10_$\rho$ | -1.34 | 3.3e-2 | 0.87 | normal | estimated |
| rate constant for conversion of target cells to a refractory state (log_10_ cell^-1^day^-1^ ) | log_10_$\phi$ | -4.99 | 3.8e-2 | 1.13 | normal | estimated |
| infected cell clearance rate (day^-1^ cells^-1^) | $\delta$ | 1.72 | 3.7e-2 | 0.54 | lognormal | estimated |
| onset of acquired immunity relative to detection (days) | $\tau$ | 6.62 | 0.32 | 0.42 | lognormal | estimated |
| Deviation from $\tau$ for unvaccinated/no record | $\beta_{\tau\_>1vax}$ | -0.032 | 3.6e-2 | -- | -- | -- |
| Deviation from $\tau$ for NBA Omicron Individuals | $\beta_{\tau_{PLAT,PAN}}$ | 0.39 | 3.6e-2 | -- | -- | -- |
| Increase in clearance rate of infected cells due to acquired immunity (day^-1^) | $m$ | 19.51 | 0.84 | 0.6 | lognormal | estimated |
| time of infection relative to first detection (days) | $t_{0}$ | 3.89 | 0.13 | 0.44 | logit[0,20] | estimated |
| Deviation from $t_{0}$ for NBA | $\beta_{t_{0},PLAT,PAN}$ | -0.71 | 4.4e-2 | -- | -- | -- |
| Potency reduction factor for PLATCOV treatment arm | log_10_paf | -0.93 | 0.071 | 0.41 | lognormal | estimated |
| initial viral inoculum (RNAcopies/mL) | $V_{0}$ | 97 |  | -- | -- | fixed |
| viral clearance rate (day^-1^) | $\gamma$ | 15 |  | -- | -- | Goyal et al. |
| mean eclipse phase duration (days^-1^) | $1/k$ | 1/4 |  | -- | -- | Ke et al. |
| Initial number of susceptible cells | $S_{0}$ | $1 \times{10}^{7}$ |  | -- | -- | Ortiz et al. |
| Initial number of refractory cells | $R_{0}$ | 0 |  | -- | -- | -- |
| Initial number of productively infected cells | $I_{P,0}$ | 0 |  | -- | -- | -- |
| Initial number of infected cells in eclipse phase | $I_{E,0}$ | 0 |  | -- | -- | -- |

**Table S4. Population parameters of combined viral dynamics + PKPD model fit to PANORAMIC+PLATCOV + NBA Omicron data*.*** Using the viral dynamic+PKPD model we estimated model parameters for Omicron infections in the NBA cohort and control and treatment arms of PLATCOV trial. For the mixed-effect model, a constant error model was used, and the magnitude of measurement error was fixed at a = 0.4 log10 copies viral RNA/ml. Vaccine status (0 vs >1 dose) was set as a covariate on $\tau$, and the cohort (NBA vs PLATCOV and PANORAMIC) was set as a covariate for $\tau$ and $t_{0}$. Also, linear dependencies between ($\pi, \phi, \delta)$ and ($\tau, \rho$) were set up in Monolix. The population parameters are recorded here and estimated individual parameters are available at https://github.com/sEsmaeili/MolnupiravirModeling.
